# Indirect effects of the first two years of the COVID-19 pandemic on secondary care for cardiovascular disease in the UK: an electronic health record analysis across three countries

**DOI:** 10.1101/2022.10.13.22281031

**Authors:** F Lucy Wright, Kate Cheema, Raph Goldacre, Nick Hall, Naomi Herz, Nazrul Islam, Zainab Karim, David Moreno-Martos, Daniel R Morales, Daniel O’Connell, Enti Spata, Ashley Akbari, Mark Ashworth, Mark Barber, Norman Briffa, Dexter Canoy, Spiros Denaxas, Kamlesh Khunti, Amanj Kurdi, Mamas Mamas, Rouven Priedon, Cathie Sudlow, Eva JA Morris, Ben Lacey, Amitava Banerjee, the CVD-COVID-UK Consortium

## Abstract

**Background:** Although morbidity and mortality from COVID-19 have been widely reported, the indirect effects of the pandemic beyond 2020 on other major diseases and health service activity have not been well described.

**Methods:** Analyses used national administrative electronic hospital records in England, Scotland and Wales for 2016-2021. Admissions and procedures during the pandemic (2020-2021) related to six major cardiovascular conditions (acute coronary syndrome, heart failure, stroke/transient ischaemic attack, peripheral arterial disease, aortic aneurysm, and venous thromboembolism) were compared to the annual average in the pre-pandemic period (2016-2019). Differences were assessed by time period and urgency of care.

**Results:** In 2020, there were 31,064 (−6%) fewer hospital admissions (14,506 [-4%] fewer emergencies, 16,560 [-23%] fewer elective admissions) compared to 2016-2019 for the six major cardiovascular diseases combined. The proportional reduction in admissions was similar in all three countries. Overall, hospital admissions returned to pre-pandemic levels in 2021. Elective admissions remained substantially below expected levels for almost all conditions in all three countries (−10,996 [-15%] fewer admissions). However, these reductions were offset by higher than expected total emergency admissions (+25,878 [+6%] higher admissions), notably for heart failure and stroke in England, and for venous thromboembolism in all three countries. Analyses for procedures showed similar temporal variations to admissions.

**Conclusion:** This study highlights increasing emergency cardiovascular admissions as a result of the pandemic, in the context of a substantial and sustained reduction in elective admissions and procedures. This is likely to increase further the demands on cardiovascular services over the coming years.

**Key Question:** What is the impact in 2020 and 2021 of the COVID-19 pandemic on hospital admissions and procedures for six major cardiovascular diseases in England, Scotland and Wales?

**Key Finding:** In 2020, there were 6% fewer hospital admissions (emergency: -4%, elective: -23%) compared to 2016-2019 for six major cardiovascular diseases, across three UK countries. Overall, admissions returned to pre-pandemic levels in 2021, but elective admissions remained below expected levels.

**Take-home Message:** There was increasing emergency cardiovascular admissions as a result of the pandemic, with substantial and sustained reduction in elective admissions and procedures. This is likely to increase further the demands on cardiovascular services over the coming years.

## Introduction

Since the early stages of the coronavirus (COVID-19) pandemic, acute workload on health systems managing those with the virus has led to direct effects (e.g. hospitalisations, intensive care admissions and mortality of infected individuals)(1–3). In addition, indirect effects have impacted non-COVID diseases by health system strain and changes in behaviours, documented across some individual specialties, clinical procedures, and countries but only in the first year of the pandemic(4–7).

The role of non-communicable diseases (NCDs) and their care as a COVID-related risk factor and outcome is consistent with a “syndemic”, “characterised by biological and social interactions between conditions and states, interactions that increase a person’s susceptibility to harm or worsen health outcomes” (8). However, pandemic planning and preparedness excludes modelling of indirect effects, which have not been of this scale in prior public health emergencies. Moreover, pandemic monitoring has focused on metrics of infection, excluding NCDs as a risk factor or indirect outcome(9).

Cardiovascular disease (CVD) is the greatest burden of disease in UK and globally(10). Any attempt to quantify indirect effects or NCDs must consider CVD. Even in the early first wave, there were indirect effects across 6 CVD subtypes in 9 UK hospitals with reduced admissions, emergency department attendances and procedures after lockdown (23 March 2020) by 58%, 53%, 31%-88% respectively, compared with prior years(2). Several studies confirmed indirect effects in different CVD subtypes and countries(7, 11-13). However, over two years into the pandemic with increasing non-COVID care backlogs in the UK and other countries, three questions remain. First, “Is the risk profile of individuals with CVD different before and during COVID-19?”, which could inform risk prediction models and CVD prevention priorities during pandemics. Second, “How has clinical activity varied across subtypes(14), admissions and procedures during the pandemic?”, to understand impact of changing pandemic waves and policy landscapes, including vaccinations and lockdowns. Third, “Are CVD admissions and procedures affected more for elective or emergency activity?”, to inform service planning and resource utilisation during and post-COVID-19. In the <u>CVD-COVID-UK/COVID-IMPACT consortium</u>, national electronic health record (EHR) data are available for pandemic-related research(15,16).

## Objectives

Using EHR phenotypes for CVD and associated procedures(14), the indirect impact of the COVID pandemic on CVD can be studied with access to data for 65.7 million individuals across multiple sources with >700 validated phenotyping algorithms. For six major CVD subtypes (acute coronary syndrome, heart failure, stroke/transient ischaemic attack, peripheral arterial disease, aortic aneurysm, and venous thromboembolism) in three UK countries (England, Scotland and Wales), we investigated hospital activity before (2016 to 2019) and during the COVID-19 pandemic (2020 to 2021) by: (i) demographic characteristics; (ii) admissions and procedures; and (iii) urgency of care.

## Methods

### Setting and data sources

National administrative hospital records for England, Scotland and Wales were used for this study and data sources are shown in **Figure S1**. Data were accessed through each country’s trusted research environment (TRE), which was made possible through agreements with Health Data Research UK for the British Heart Foundation Data Science Centre’s CVD-COVID-UK/COVID-IMPACT research programme (15, 16). For England, data were obtained from the Admitted Patient Care Hospital Episode Statistics in NHS Digital’s TRE service for England. For Scotland, the Scottish Morbidity Records (SMR 01) for General / Acute Inpatient and Day Case admission in the Scottish National Safe Haven was the data source (17) and for Wales, it was the Patient Episode Database Wales in the SAIL Databank (18). These datasets cover inpatient admissions to all NHS hospitals including day cases.

Data extraction and analysis of patient-level hospital data was undertaken in each nation’s TRE using common data specifications and analysis codes, accounting for differences in data structure and clinical coding procedures between the three nations. In the raw form, each record represents a new admission, a change between medical specialists within the same admission or an interhospital transfer. To minimise overcounting, a record that represented a continuous hospital stay and included changes between medical specialists within the same admission and accounted for interhospital transfers was created. Only aggregated data were shared.

### Study population

The study population included all individuals admitted to hospital in England, Scotland or Wales with a primary diagnosis of each CVD subtype between January 1^st^, 2016 and December 31^st^, 2021– the study period covers four years before the COVID-19 pandemic for comparison with the first two years of the pandemic. The study population also included all individuals admitted for each of the associated CVD procedures to ensure that all procedures were captured, since the associated CVD subtype diagnosis might not necessarily be recorded as the primary diagnosis for these admissions.

### Admissions for CVD diagnoses and procedures

The CVD diagnoses included in this study were acute coronary syndrome (ACS), heart failure (HF), acute stroke or transient ischaemic attack (stroke/TIA), peripheral arterial disease (PAD), aortic aneurysm (AA) and venous thromboembolism (VTE). The associated CVD procedures were: percutaneous coronary intervention, coronary artery bypass graft surgery, pacemaker or cardiac resynchronisation therapy, ventricular assist device or heart transplant, stroke thrombolysis or thrombectomy, carotid endarterectomy or stenting, cerebral aneurysm coiling, aortic aneurysm repair, peripheral limb angioplasty, limb revascularisation, bypass or amputation, and pulmonary artery embolectomy or embolisation.

Phenotypes for CVD diagnoses were defined using the international classification of diseases, 10th revision (ICD-10 codes) and procedures using office of population censuses and surveys classification of interventions and procedures, version 4 (OPCS-4 codes). These were chosen to align with an earlier study (2) and a few minor modifications were made to OPCS-4 codes after clinical and academic expert consensus (e.g. additional codes for coronary artery bypass surgery or pacemaker insertion). For procedures, we counted all recorded procedures in a single admission. Details of diagnostic and procedural clinical terminology codes are in **Table S1**.

Admissions, not individuals, were counted. One patient may have been readmitted for a CVD subtype or procedure, or had an admission for other CVD subtypes or procedures during the study period, and each admission was counted. Admissions were classified as emergency or elective using the admission type variable in each dataset. Generally, emergency admissions were when the admission was unpredictable and at short notice due to clinical need, such referral from accident and emergency, a general practitioner or a clinic. Admissions were classified as elective when the decision to admit could be separated from the time of the actual admission such as being admitted from a waiting list, or having the admission booked or planned at the time when it was deemed clinically necessary.

### Statistical analysis

Analyses were for men and women combined and for all ages. Demographic characteristics of individuals admitted to hospital pre-pandemic with those admitted during the first two years of the pandemic for CVD subtypes and procedures were investigated. The characteristics were for each admission and included sex, age, and ethnic group, and Charlson comorbidity index using ICD-10 codes from the admission(19).The average number of people in each category of the selected demographic characteristic was divided by the average number of admissions. Due to small numbers (<5) in some categories, characteristics for 2020 and 2021 were reported as an average of the two years. For all other analyses, 2020 and 2021 are reported separately. The average of 2016-2019 was selected as the comparator for all analyses, using a four-year period to give stability over time.

Initially all admissions (emergency and elective combined) were assessed. To explore indirect effects of the pandemic on unplanned and planned CVD care and inform policy responses, admissions were reported separately as emergency or elective, respectively on CVD admissions to hospital separately from hospital capacity to provide planned care during the pandemic. Annual counts were calculated within the three time periods of interest: pre-pandemic 2016-2019, and pandemic 2020 and 2021. The percentage change between the time periods was calculated by subtracting the total for each pandemic year from the average of 2016-2019 and dividing it by the 2016-2019 average. Percentage changes were calculated with 95% confidence intervals, assuming the pre-pandemic annual counts followed a negative binomial distribution. Monthly counts of admissions were also calculated and plotted to show trends during the years. The analyses were performed according to a pre-specified protocol and analysis plan with phenotyping and analysis code, which is available at https://github.com/BHFDSC/CCU003_04.

## Results

### Study population

We identified a total of 1,973,104 and 970,374 admissions and 1,616,550 and 635,187 procedures in 2016-2019 and 2020-2021 respectively.

### Demographic characteristics

There were no major differences by age, gender or ethnic group between 2016-2019 and 2020-2021 for admissions (**Table S2**) or procedures (**Table S3**) across countries or CVD subtypes. The Charlson comorbidity profile was more severe in 2020-2021, compared with 2016-2019, for all admissions and all procedures in England, except VAD/transplant and PA embolectomy. In Scotland, individuals with PAD (20.4% vs 17.9%) and VTE (18.8% vs 17.9%) had more severe comorbidities in 2020-2021, compared with pre-pandemic, but otherwise, in Scotland and Wales, there were no notable differences by comorbidities in admissions or procedures between pre- and post-pandemic periods.

### Hospital admissions

#### Total admissions

In 2020, there were 31,064 (−6%) fewer admissions for all six CVD subtypes combined in the three countries combined compared with the expected number from 2016-2019. In 2021, there was an overall increase of 14,884 (+3%) admissions. **Figure 1** shows the annual counts and percentage change in total admissions for CVD as the primary diagnosis for all CVD and across subtypes for each country. In 2020, admissions for all CVD in the three individual countries were lower than expected (−6% in England, -6% in Scotland, -7% in Wales). In 2021, admissions in England were 4% higher than expected, but in Scotland and Wales, the numbers were similar to 2016-2019.

**Figure 1.**
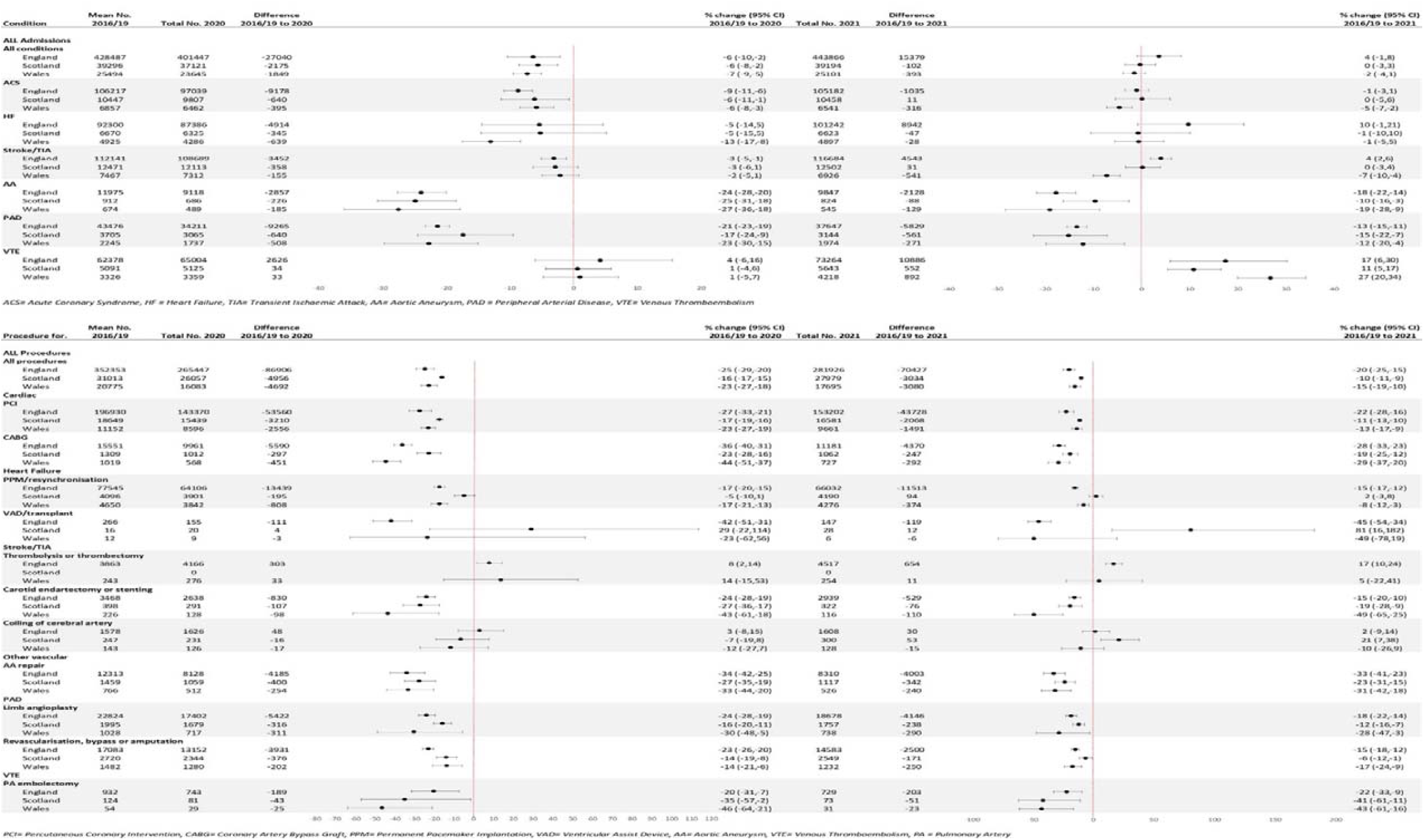
Annual counts and percentage change in total admissions and procedures between pre-pandemic (2016-2019) and pandemic (2020-2021) periods for cardiovascular disease as primary diagnosis for all cardiovascular disease and across subtypes and across three countries in the UK.

For most CVD subtypes, admissions in 2020 were lower than expected across countries compared with 2016-2019 (**Figure 1**). Admissions for ACS, AA and PAD were lower in all three countries (annual % change range: -6% to -27%), and for HF in Wales (−13%). For stroke/TIA in England, admission numbers were somewhat lower (−3%). For the remaining CVD subtypes, the observed numbers of admissions in 2020 were similar to those in 2016-2019. In 2021 in all three countries, admissions for AA and PAD continued to remain lower than expected (range: -10% AA in Scotland to -19% AA in Wales) and there were more admissions for VTE than expected (range: +11% in Scotland to +27% in Wales). In England, there were somewhat more admissions for stroke/TIA in England (+4%). In Scotland, admissions for ACS, HF and stroke/TIA were similar in 2021 compared to 2016-2019. In Wales, admissions for ACS and stroke/TIA were lower, -5% and -7% respectively.

The observed changes were not uniform throughout the two pandemic years (**Figure 2**). In 2020, monthly admissions for all CVD subtypes in all three countries decreased from January, with greatest reductions in April, compared to 2016-2019. Admissions remained lower than expected during the rest of 2020, except VTE admissions which increased above 2016-2019 levels by May/June 2020 and throughout 2021. In 2021, monthly admissions for all CVD subtypes were lower than 2016-2019 levels in January and February in all 3 countries, except VTE. Timing and extent of recovery to expected levels varied by CVD subtype and country.

**Figure 2.**
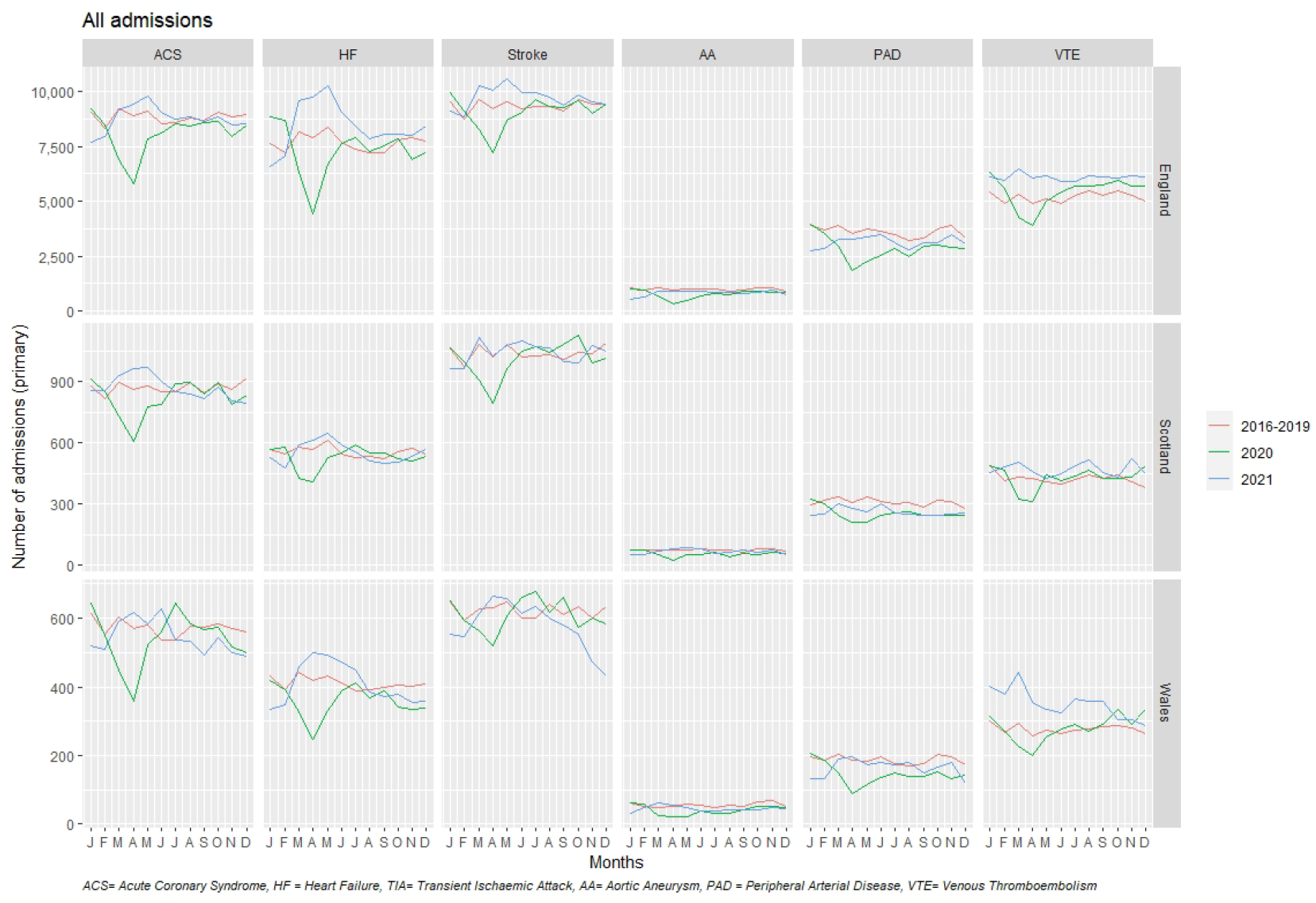
Monthly total admissions for cardiovascular disease as primary diagnosis across subtypes, across three countries in the UK and across pre-pandemic (2016-2019) and pandemic (2020-2021) periods.

#### Urgency of Care

In 2020, there were 14,506 (−4%) fewer emergency admissions than expected for all CVD in the three countries combined, and the proportion reduction was similar in all three countries. For most CVD subtypes, emergency admissions in all three countries were similar to those in 2016-2019 (**Figure 3**). The exceptions were ACS in England (−8%), HF in Wales (−13%), stroke/TIA in England (−3%), AA in England and Scotland (−9% and -12% respectively) and PAD in England (−5%). There were also fewer elective admissions for all CVD in 2020 with a total of 16,560 (−23%) fewer in the three countries (−22% for England and Wales, -30% for Scotland). For most CVD subtypes across countries compared to 2016-2019, elective admissions were lower (e.g: -16% ACS in England and stroke/TIA in Scotland, - 35% HF in Scotland). Admissions with the greatest reductions were AA (−34%, -38% and -37%) and PAD (−30%, -36% and -40%) in England, Scotland and Wales respectively. Admission numbers were similar to 2016-2019 for ACS in Scotland and Wales, Stroke/TIA in England and Wales and VTE in England and Scotland. No admissions for CVD subtypes were higher than expected.

**Figure 3:**
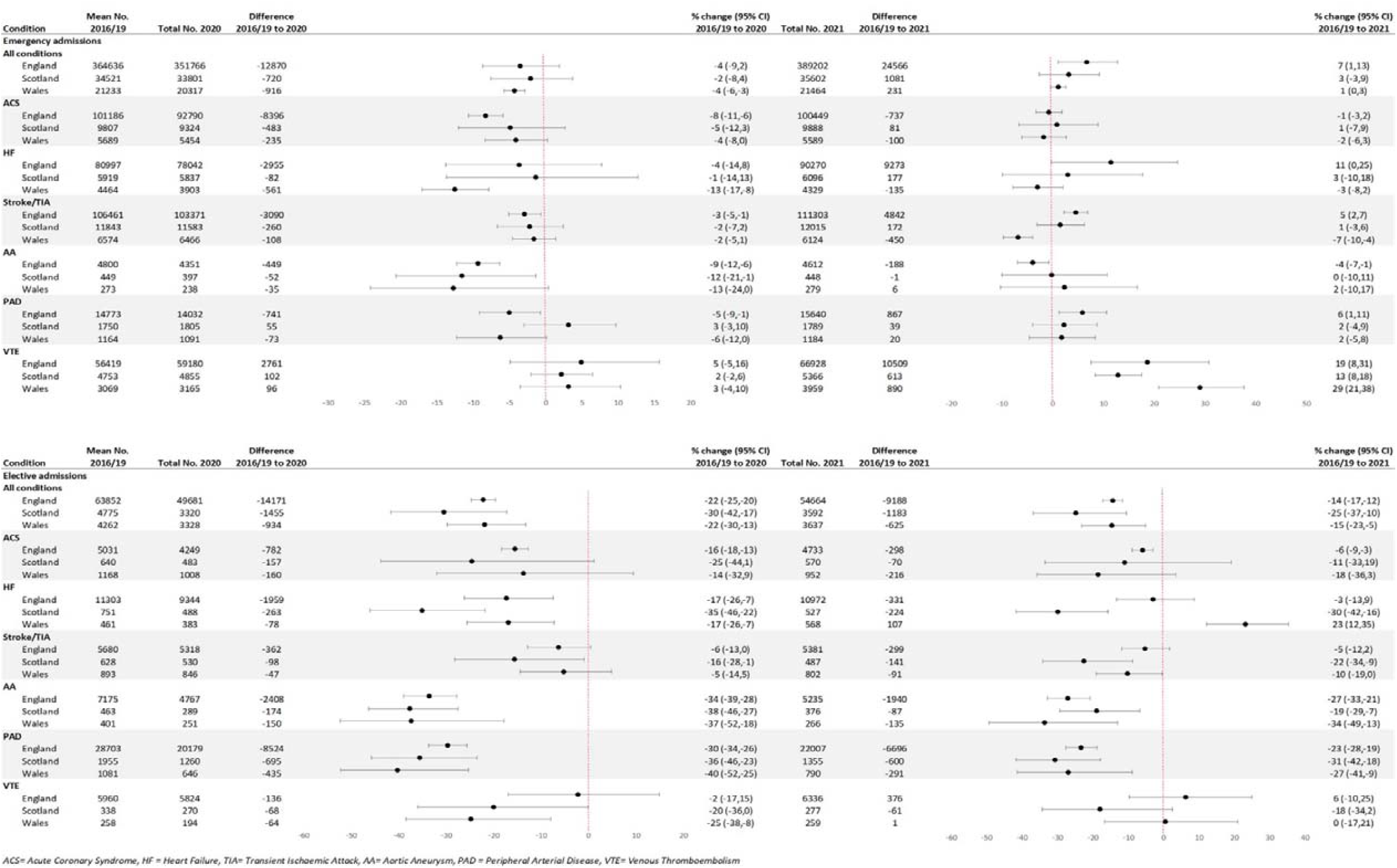
Annual counts and percentage change in total emergency and elective admissions between pre-pandemic (2016-2019) and pandemic (2020-2021) periods for cardiovascular disease as primary diagnosis for all cardiovascular disease and across subtypes and across three countries in the UK.

In 2021, there were 25,878 (+6%) more emergency CVD admissions in the three countries combined. This was driven by the 7% higher number of admissions in England, while in Scotland and Wales numbers returned to expected levels (**Figure 3**). For most CVD subtypes across countries, the number of emergency admissions returned to expected levels, although there were some exceptions. Admissions were higher than expected in all three countries for VTE (England +19%, Scotland +29%, Wales +13%). In England, admissions were higher than expected for HF (+11%), stroke/TIA (+5%) and PAD (+6%) and somewhat lower for AA (−4%). Elective admissions remained lower in 2021 with 10,996 (−15%) fewer CVD admissions than in 2016-2019 in the three countries combined (England -14%, Scotland -25%, Wales -15%). For individual CVD subtypes, elective admissions remained below pre-pandemic levels across all subtypes and countries, except HF in Wales (+23%) (**Figure 3**). Monthly emergency and elective admissions across CVD subtypes and across countries decreased between January and April 2020 (**Figures 4 and 5** respectively). Emergency activity returned to 2016-2019 levels by June/July 2020, except for VTE which remained higher throughout the rest of 2020 and 2021. Between February and April 2021, emergency CVD admissions exceeded 2016-2019 levels, then decreased, particularly in Wales, where ACS, HF and stroke admissions decreased to lower than pre-pandemic levels (**Figure 4**). Other than VTE, elective admissions did not return to expected levels by end of 2021, across CVD subtypes and countries (**Figure 5**). Reductions for elective and emergency admissions were greater in England and Wales.

**Figure 4:**
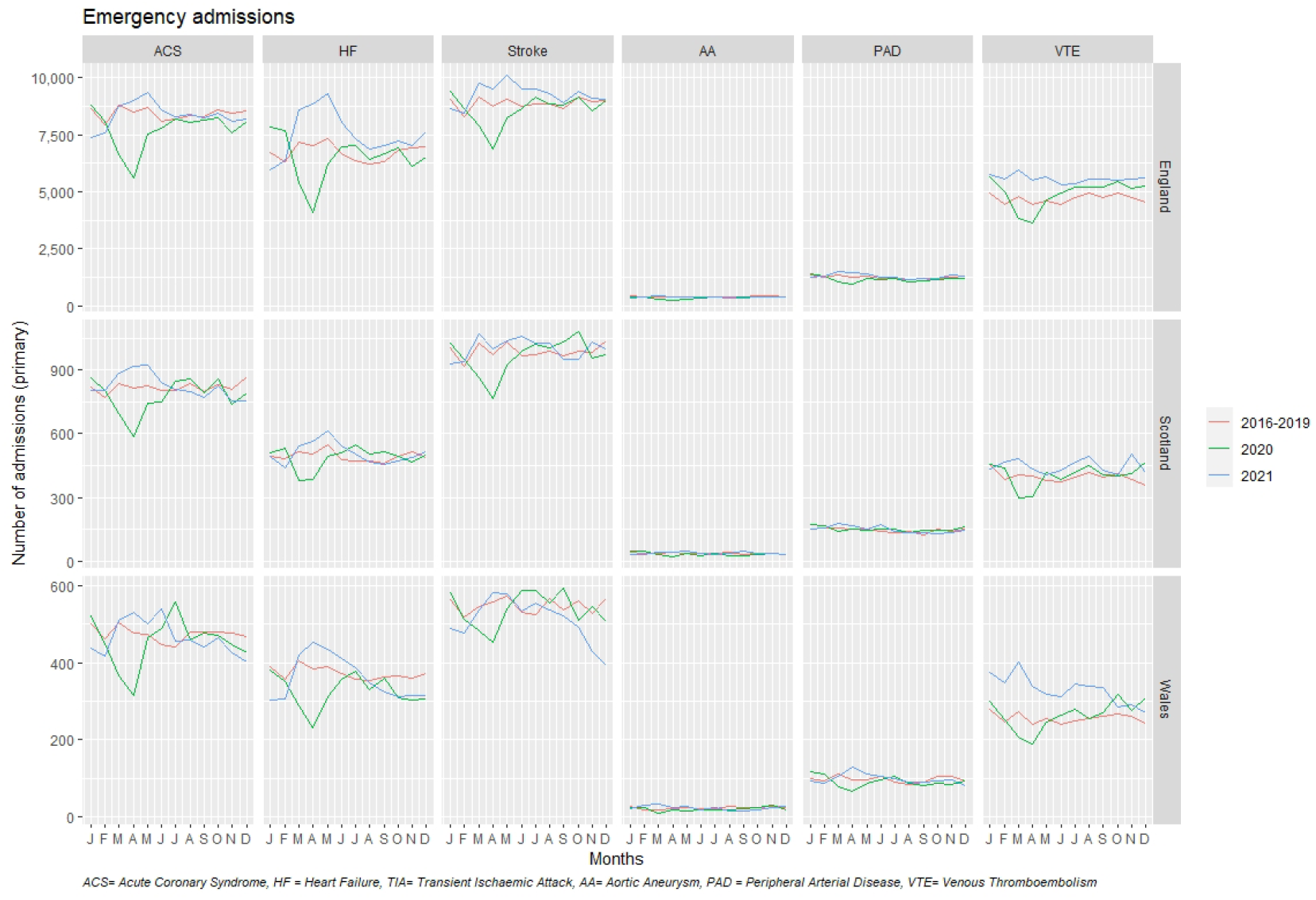
Monthly emergency hospital admissions for cardiovascular disease as primary diagnosis across subtypes, across three countries in the UK and across pre-pandemic (2016-2019) and pandemic (2020 and 2021) periods.

**Figure 5:**
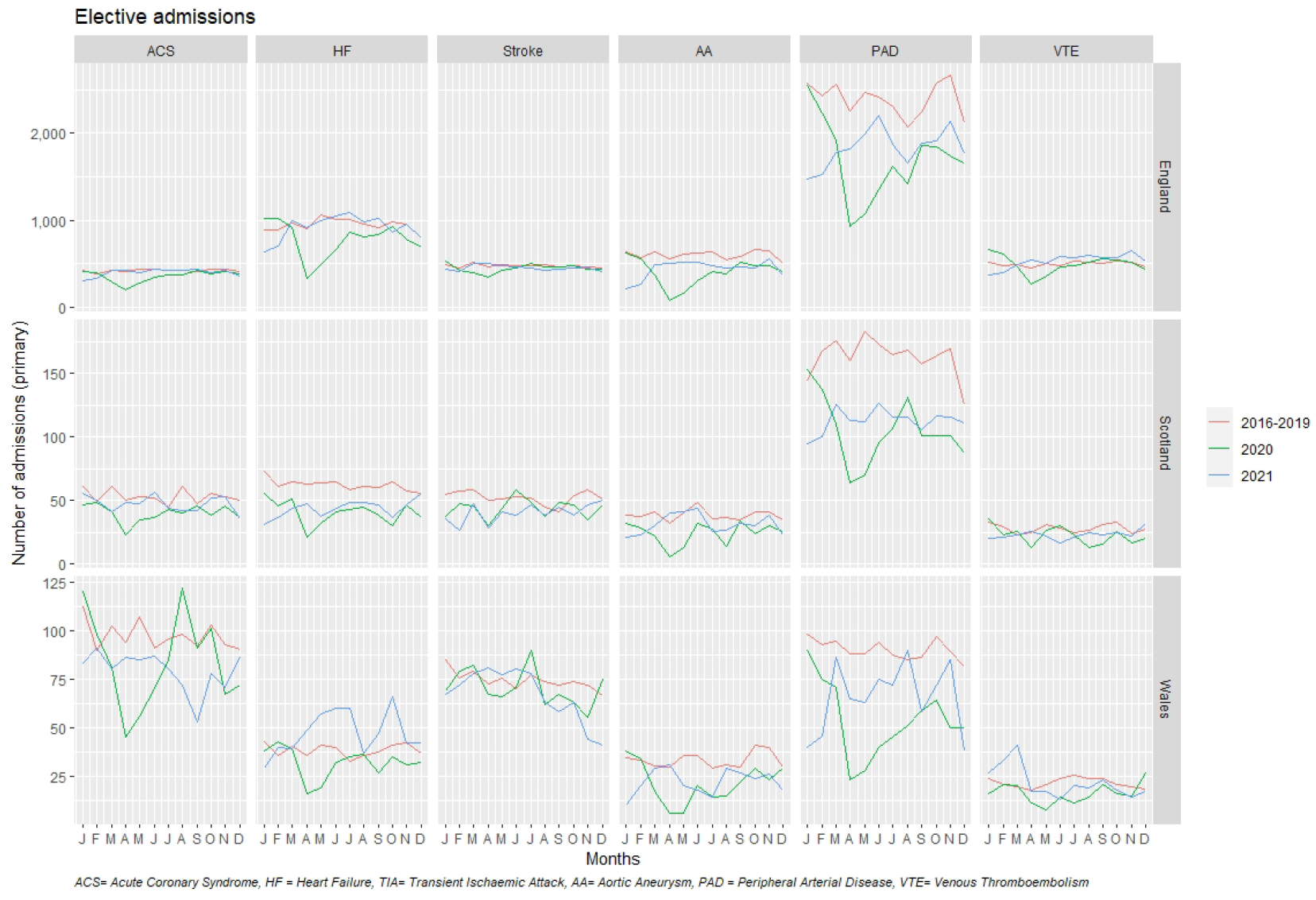
Monthly elective hospital admissions for cardiovascular disease as primary diagnosis across subtypes, across three countries in the UK and across pre-pandemic (2016-2019) and pandemic (2020-2021) periods.

### Procedures

#### Total procedures

In 2020, there were 96,554 (−24%) fewer total procedures for all six CVD subtypes combined in the three UK countries compared with the expected number in 2016-2019. In 2021, there were 76,541 (−19%) fewer CVD procedures. In 2020, admissions for all CVD procedures in the three individual countries were lower than expected and varied by country (England -25%, Scotland -16%, Wales - 23%) (**Figure 1**). In 2021, there was a small increase (5-8%) in all CVD procedures in the three countries, but numbers remained below expected levels (England -20%, Scotland -10%, Wales -15%).

There were major reductions across most individual CVD procedures in 2020 compared with 2016-2019. These included percutaneous coronary intervention (range: -17% in Scotland to -27% in England), coronary artery bypass graft surgery (−23% in Scotland to -44% in Wales), carotid endarterectomy (England -24% to Wales -43%) and limb angioplasty (−16% in England to -30% in Wales). Only stroke thrombolysis in England was higher than expected (+8%), and cerebral artery coiling in all three countries was similar to 2016-2019. In 2021, although there was some improvement, most CVD procedures remained well below the expected levels in all three countries, ranging from -6% for PAD revascularisation in Scotland to -49% for carotid endarterectomy in Wales. Only stroke thrombolysis in England (+17%), cerebral artery coiling in Scotland (+21%), and ventricular assist device or heart transplant in Scotland (+81%) were higher, but numbers were low and confidence intervals wide. Generally, monthly numbers of CVD procedures were lower in 2020 and 2021, compared to 2016-2019 across countries (**Figure 3**).

The observed changes were not consistent throughout 2020 and 2021 (**Figure 6**). In 2020, monthly admissions for all CVD procedures in all three countries decreased from January, with greatest reductions in April, compared to 2016-2019. Overall admissions for procedures all CVD subtypes remained lower than expected during the rest of 2020 and 2021 in all three countries.

**Figure 6.**
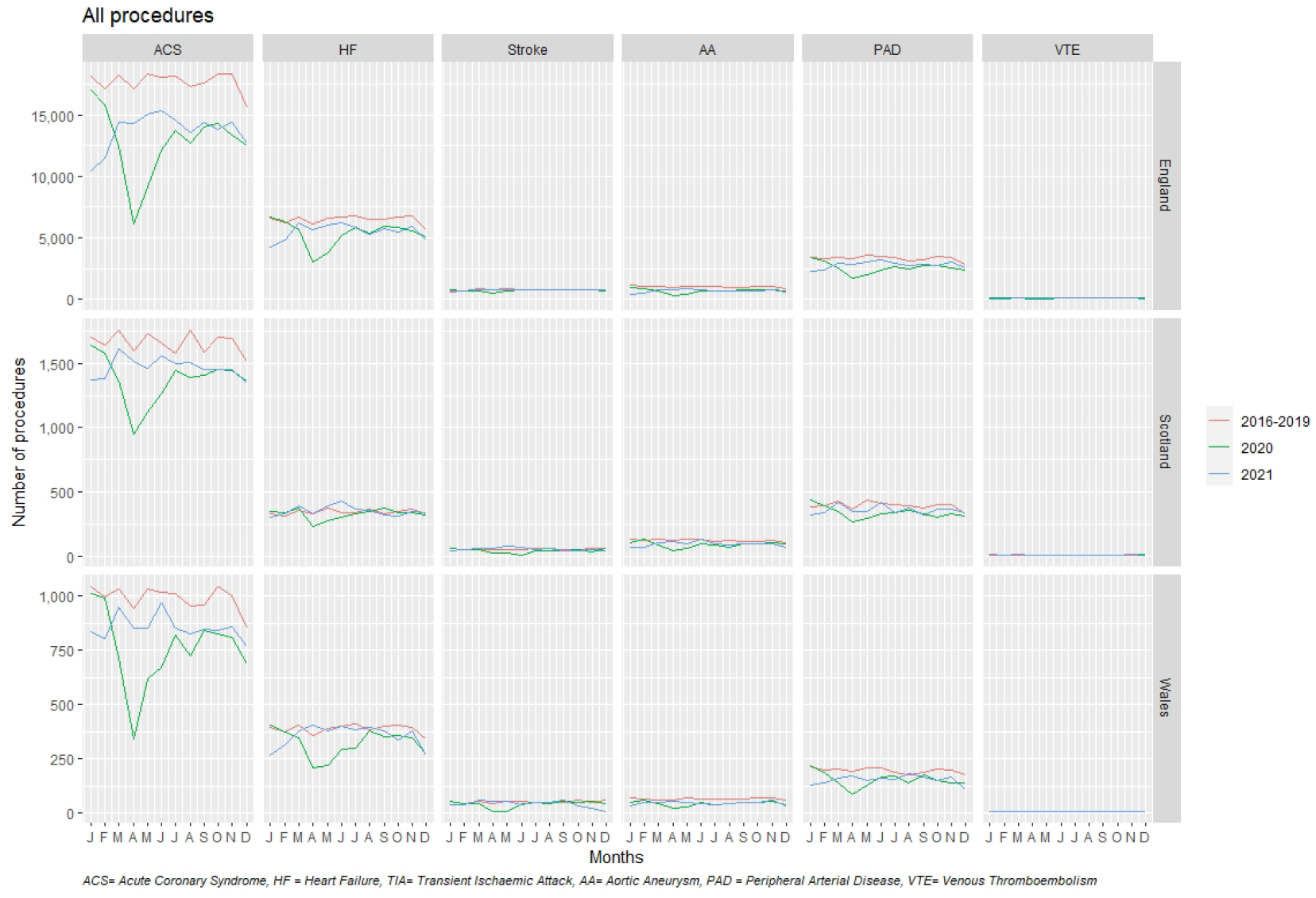
Monthly total procedures for cardiovascular disease across subtypes, across three countries in the UK and across pre-pandemic (2016-2019) and pandemic (2020-2021) periods.

#### Urgency of Care

In 2020 there were 11,775 (−9%) fewer emergency CVD procedures than in 2016-2019 in all three countries combined. The proportion reduction varied between countries (England -10%, Scotland +3%, Wales -4%) (**Figure 7**). The total number of individual emergency CVD procedures was either lower than or similar to 2016-2019. Examples of procedures that were lower were coronary artery bypass graft surgery (−26% in England, -57% in Wales), carotid endarterectomy (−36% in Wales), AA repair (−23% in England, -25% in Wales) and pulmonary artery embolectomy (−20% in England). Some procedures were higher in 2020 than expected: permanent pacemaker or resynchronisation therapy in Scotland (+20%) and cerebral artery coiling (+15% in England and +47% in Wales), although numbers were relatively low.

**Figure 7:**
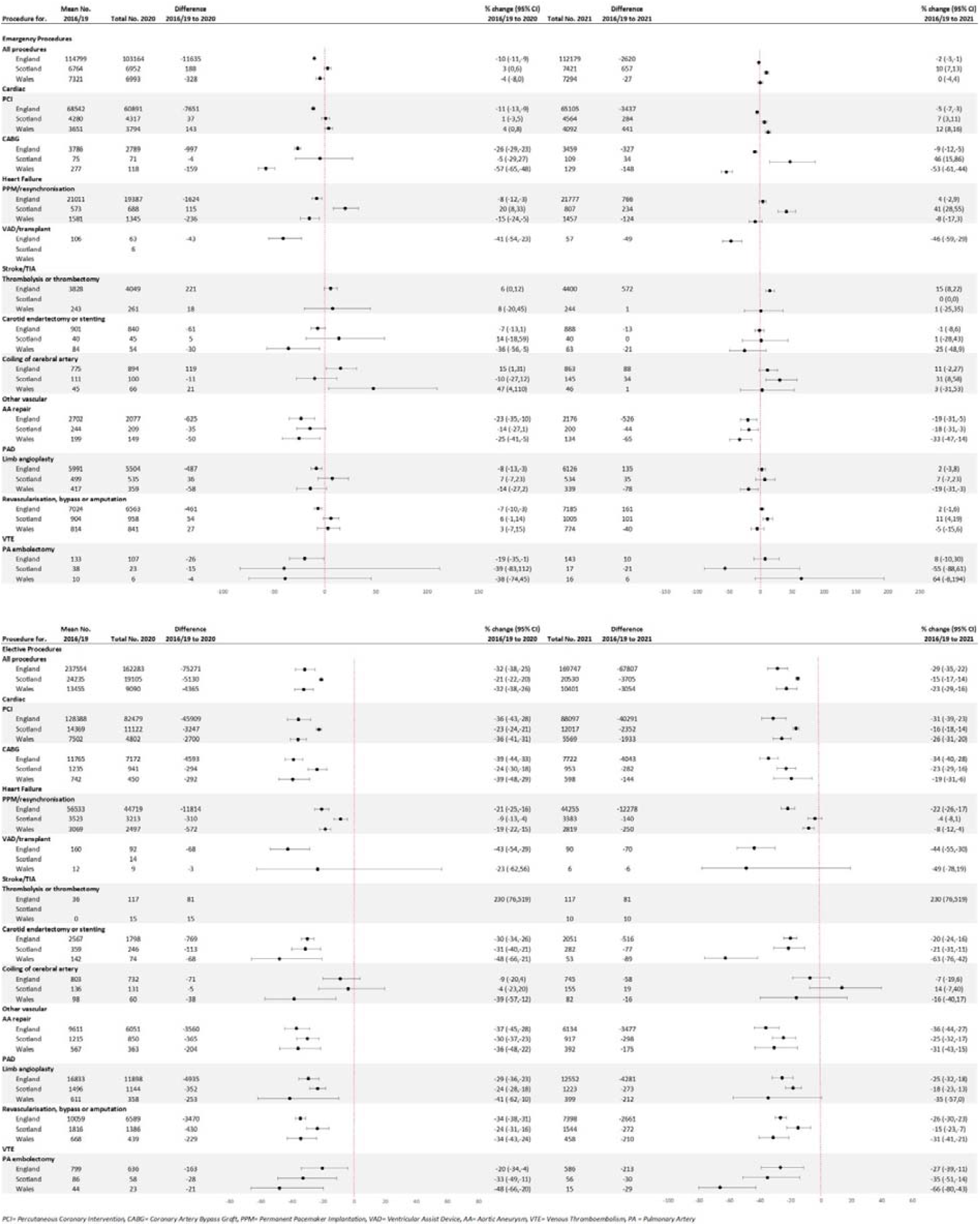
Annual counts and percentage change in total emergency and elective procedures between pre-pandemic (2016-2019) and pandemic (2020-2021) periods for cardiovascular disease for all cardiovascular procedures and across subtypes and across three countries in the UK.

In 2021 there were only 1,990 (−2%) fewer emergency CVD procedures in all three countries combined, with some variation observed between countries (England -2%, Scotland +10%, Wales 0%). Generally, there was variability across individual CVD procedures and countries (**Figure 7**). For example, emergency coronary artery bypass graft surgery was lower in England (−9%) and Wales (−53%) and higher in Scotland (+46%). However, AA repair across all three countries remained lower in 2021 than 2016-2019 (England -19%, Scotland -18%, Wales -33%).

For elective CVD procedures in 2020, there were 84,766 (−31%) fewer procedures combined for all three countries (England and Wales -32%, Scotland -21%). Individual elective procedures were all lower in 2020 compared to 2016-2019 across countries, except stroke thrombolysis was higher in England (+230%, N+81). The reduction in CVD procedures varied by country and procedure, e.g. - 39%, -39% and -24% for CABG, and -29%, -24% and -41% for limb angioplasty in England, Wales and Scotland, respectively.

In 2021, the reduction in elective procedures persisted with 74,566 (−27%) fewer elective CVD procedures combined for all three countries (England -29%, Scotland -15%, Wales -23%) (**Figure 7**). The reductions continued for all individual elective procedures, except stroke thrombolysis in England. Between January and April 2020, monthly emergency procedures decreased for AA, ACS, HF and PAD, recovering to pre-pandemic levels in late 2020 and 2021. (**Figure S2**). Monthly elective procedures decreased in January-April 2020, across subtypes and countries, and had not recovered to 2016-19 levels by end of 2021 (**Figure S3**).

## Discussion

In the first comprehensive study to use national routinely collected electronic hospital data in the pandemic context across CVD subtypes, admissions, procedures, urgency of care and countries, we demonstrate three major findings. First, there were profound reductions across CVD subtypes and countries during the pandemic, particularly for procedural activity which reduced by a third in 2020 and by a quarter in 2021, compared with pre-pandemic levels. Second, except for VTE, although emergency admissions and procedures had returned to pre-pandemic levels by 2021, elective activity remained significantly reduced, especially for procedures. Third, the comorbidity profile for CVD admissions and procedures was more severe during the pandemic than pre-pandemic in England for most CVD subtypes but did not generally differ between 2020-21 and 2016-19 for Scotland and Wales.

Despite multiple analyses of indirect effects in the UK and other countries using EHR(11-14, 20), these effects have been neglected in pandemic surveillance and policy responses(21). Moreover, prior analyses have tended to be disease- or procedure-specific and have not taken a system-level view across diseases and countries(22). We now confirm previous reports of reduced activity for admissions and for PCI and other CVD-related procedures, showing variation by timing, speed and extent of recovery across subtypes. Given the significant backlogs across services in the UK(23-25) and other countries(26), there is an urgent need to monitor and understand these indirect impacts of the COVID-19 pandemic, to develop coordinated, but tailored responses, based on subtype, type and urgency of care, and country. Without urgent action, indirect and long-term consequences could create far greater burden and cost to individuals, populations and health systems than acute, direct effects.

Reductions in emergency care are likely to require different approaches to workforce and resource planning, compared with elective care, and for admissions versus procedures(27, 28). Therefore, the greater effect on procedural activity and the relatively slower recovery of elective procedural activity, especially in England and Wales, requires further investigation, explanation and mitigation strategies. The widespread strain on health systems due to COVID-19 is unprecedented and staff and resource shortages over successive waves may provide part of the explanation. In 2021, some emergency admissions were greater than pre-pandemic levels, which may in part, be related to the reduction in elective admissions and procedures the year before. There may also have been changes in coding of admissions as “emergency” or “urgent” so that they were less delayed during the pandemic.

Although projections from national and international efforts such as the Global Burden of Disease Study provide important context(29), more detailed national-and local-level data are required for informed health policy. Ultimately, answers require standardised, near real-time data which has become possible in the COVID-19 context but has historically not been a priority. To-date, admissions and procedures have been tracked in a “rear view mirror”, which is not fit-for-purpose for surveillance and planning during public health emergencies such as pandemics, due partly to specialty- and disease-specific silos, and partly due to a culture of retrospective data collection, monitoring and analysis. For example, the UK’s National Heart Failure Audit and National Audit of PCI publish annual reports with a one year delay, which has been further delayed or de-prioritised during the pandemic(30) and national AA screening data during the pandemic has not been published(31). National EHR data can and should be used to study CVD and non-COVID diseases and services at scale, with low-hanging fruits for public health and policy planning during and post-pandemic.

Late presentation, greater severity of illness and inequalities in access to healthcare have been invoked to explain increased rates of CVD during the pandemic. The finding of similar baseline characteristics before and during the pandemic among most individuals presenting with CVD and undergoing CVD-related procedures in Scotland and Wales suggests that these patient-level factors (including age, sex and ethnicity) do not fully explain the reductions in CVD care during the pandemic, and that system-level factors may be more important. However, for most CVD subtypes in England, there was greater comorbidity burden in those presenting during pandemic years than during pre-pandemic years, which could suggest decreased prevention, late presentation and/or reduced access to CVD services. We show differences by subtype, by urgency of care and by country, which may signify different reasons for reductions in activity and therefore different, nuanced solutions. For example, the higher impact on CABG, carotid endarterectomy and PAD procedures needs to be explored. There is now clear evidence of increased VTE risk associated with COVID-19, up to 1 year after infection, which at least partly explains the observed increase in VTE admissions and procedures(32). We have only considered certain CVD admissions and procedures, but indirect effects across all diseases and procedures are likely. A complex interplay of factors makes analysis difficult with successive pandemic waves, lockdowns, vaccination programmes and changing COVID-19-related policies. However, without a whole-system perspective and better up-to-date data, the indirect effect across diseases cannot be quantified, tackled or predicted. Only then can the correct, nuanced approaches to workforce, public health priorities, health resource utilisation be planned.

### Strengths/limitations

Our analyses used standardised, validated, open-source coding in national level EHR data in the most comprehensive investigation of indirect effects to-date. We used pre-pandemic data for comparison. We used the same methods for admissions and procedures across diseases, CVD subtypes and comparable datasets across countries (17). Our study does have some limitations. First, we only investigated some, not all CVD admissions and procedure. Second, there were low numbers for certain procedures, particularly in Scotland and Wales. Third, we only examined inpatient and not outpatient or emergency department activity. Fourth, we only had sociodemographic and comorbidity data at baseline and not tracked over time. Fifth, we do not look in detail at impact of lockdowns, vaccination, successive waves. Sixth, we are reliant on EHR and coding errors are possible though prior published studies suggest that this unlikely to be a major issue. Seventh, we did not investigate impact of changes in CVD admissions or procedures on CVD-related mortality, which should be considered in future studies. Finally, our analyses only concerned UK data, and may not be necessarily generalisable to other countries and settings.

### Implications for clinical practice and policy

Our results suggest that procedural activity needs to be prioritised and planned to provide timely services for high-risk patients during pandemics. Data about potential indirect effects needs to be collected and monitored, and ways of collecting, storing and analysing data need to be standardised across diseases and procedures, i.e. we cannot have every specialty and disease developing its own methods and “re-inventing the wheel”. During planning for pandemics, NCD surveillance needs to be part of the preparation and during pandemics, it should be part of the dashboards.

### Implications for research

With the advent of national TRE data, the type of research which we have conducted needs to be scaled up. Open-source data and methods can facilitate valid comparisons within and across countries, but differences in capture and coding of data need to taken into account. Our methods and our results have application to other diseases (19,20) and other countries, with potential implications for current and future pandemic policy. The science of pandemic preparedness has been largely restricted to infection dynamics. Future prediction models have to incorporate NCDs, and should include indirect effects, which may be as profound as direct effects.

## Conclusions

There have been wide and far-reaching reductions in secondary care for cardiovascular disease throughout the pandemic, with incomplete recovery, particularly for procedural and elective activity, even two years into the COVID-19 pandemic. Monitoring and protection of cardiovascular and non-COVID services should be part of pandemic planning in future.

## Data Availability

The data used in this study are available in the NHS Digital TRE for England but as restrictions apply they are not publicly available (https://digital.nhs.uk/coronavirus/coronavirus-data-services-updates/trusted-research-environment-service-for-england). The CVD-COVID-UK/COVID-IMPACT programme led by the BHF Data Science Centre (https://www.hdruk.ac.uk/helping-with-health-data/bhf-data-science-centre/) received approval to access data in NHS Digital TRE for England from the Independent Group Advising on the Release of Data (IGARD) (https://digital.nhs.uk/about-nhs-digital/corporate-information-and-documents/independent-group-advising-on-the-release-of-data) via an application made in the Data Access Request Service (DARS) Online system (ref. DARS-NIC-381078-Y9C5K) (https://digital.nhs.uk/services/data-access-request-service-dars/dars-products-and-services). The CVD-COVID-UK/COVID-IMPACT Approvals & Oversight Board (https://www.hdruk.ac.uk/projects/cvd-covid-uk-project/) subsequently granted approval to this project to access the data within NHS Digital TRE for England, the Scottish National Safe Haven and the Secure Anonymised Information Linkage (SAIL) Databank. The de-identified data used in this study were made available to accredited researchers only. Those wishing to gain access to the data should contact in the first instance.
Data used in this study are available in the Scottish National Safe Haven (Project Number: 2021-0102), but as restrictions apply they are not publicly available. Access to data may be granted on application to, and subject to approval by, the Public Benefit and Privacy Panel for Health and Social Care (PBPP (https://www.informationgovernance.scot.nhs.uk/pbpphsc/)). Applications are coordinated by eDRIS (electronic Data Research and Innovation Service (https://www.isdscotland.org/Products-and-services/Edris/)). The anonymised data used in this study was made available to accredited researchers only through the Public Health Scotland (PHS) eDRIS User Agreement (https://www.isdscotland.org/Products-and-services/Edris/_docs/eDRIS-User-Agreement-v16.pdf).
The data used in this study are available in the SAIL Databank at Swansea University, Swansea, UK, but as restrictions apply they are not publicly available. All proposals to use SAIL data are subject to review by an independent Information Governance Review Panel (IGRP). Before any data can be accessed, approval must be given by the IGRP. The IGRP gives careful consideration to each project to ensure proper and appropriate use of SAIL data. When access has been granted, it is gained through a privacy protecting safe haven and remote access system referred to as the SAIL Gateway. SAIL has established an application process to be followed by anyone who would like to access data via SAIL at https://www.saildatabank.com/application-process

## Acknowledgements

This work is carried out with the support of the BHF Data Science Centre led by HDR UK (BHF Grant no. SP/19/3/34678). This study makes use of de-identified data held in NHS Digital’s TRE for England, the SAIL Databank and the Scottish National Data Safe Haven and made available via the BHF Data Science Centre’s CVD-COVID-UK/COVID-IMPACT consortium. This work uses data provided by patients and collected by the NHS as part of their care and support. We would also like to acknowledge all data providers who make health relevant data available for research.

The study makes use of anonymised data held in the Scottish National Safe Haven. The authors would like to acknowledge the support of the eDRIS Team (Public Health Scotland) for their involvement in obtaining approvals, provisioning and linking data and the use of the secure analytical platform within the National Safe Haven.

This study makes use of anonymised data held in the Secure Anonymised Information Linkage (SAIL) Databank. This work uses data provided by patients and collected by the NHS as part of their care and support. We would also like to acknowledge all data providers who make anonymised data available for research. We wish to acknowledge the collaborative partnership that enabled acquisition and access to the de-identified data, which led to this output. The collaboration was led by the Swansea University Health Data Research UK team under the direction of the Welsh Government Technical Advisory Cell (TAC) and includes the following groups and organisations: the SAIL Databank, Administrative Data Research (ADR) Wales, Digital Health and Care Wales (DHCW), Public Health Wales, NHS Shared Services Partnership (NWSSP) and the Welsh Ambulance Service Trust (WAST). All research conducted has been completed under the permission and approval of the SAIL independent Information Governance Review Panel (IGRP) project number 0911.

The Big Data Institute has received funding from the Li Ka Shing Foundation and Robertson Foundations, the Medical Research Council, British Heart Foundation, and is supported by the NIHR Oxford Biomedical Research Centre.

## Authors contributions

AB, FLW, and EM conceptualised the study. FLW and AB were the project leads. FLW and AB created the data specifications. NHall, KC, DO’C, ZK and DM-M were responsible for data extraction. NI, KC, DO’C, D M-M and RG conducted data analyses. RG and ES provided statistical advice. FLW, EM, BL, RG and AB reviewed the methods and interpreted the data. KC, NHerz and NI produced the graphics. CS is the Director of the BHF Data Science Centre and coordinated approvals for and access to data within NHS Digital’s TRE for England, the SAIL Databank and the Scottish National Safe Haven for CVD-COVID-UK/COVID-IMPACT. FLW and AB drafted the manuscript and all authors reviewed and edited the final version of the manuscript.

## Ethics statement

The North East - Newcastle and North Tyneside 2 research ethics committee provided ethical approval for the CVD-COVID-UK/COVID-IMPACT research programme (REC No 20/NE/0161) to access, within secure trusted research environments, unconsented, whole-population, de-identified data from electronic health records collected as part of patients’ routine healthcare.

## Data availability statement

The data used in this study are available in NHS Digital’s TRE for England, but as restrictions apply they are not publicly available (https://digital.nhs.uk/coronavirus/coronavirus-data-services-updates/trusted-research-environment-service-for-england). The CVD-COVID-UK/COVID-IMPACT programme led by the BHF Data Science Centre (https://www.hdruk.ac.uk/helping-with-health-data/bhf-data-science-centre/) received approval to access data in NHS Digital’s TRE for England from the Independent Group Advising on the Release of Data (IGARD) (https://digital.nhs.uk/about-nhs-digital/corporate-information-and-documents/independent-group-advising-on-the-release-of-data) via an application made in the Data Access Request Service (DARS) Online system (ref. DARS-NIC-381078-Y9C5K) (https://digital.nhs.uk/services/data-access-request-service-dars/dars-products-and-services). The CVD-COVID-UK/COVID-IMPACT Approvals & Oversight Board (https://www.hdruk.ac.uk/projects/cvd-covid-uk-project/) subsequently granted approval to this project to access the data within NHS Digital’s TRE for England, the Scottish National Safe Haven and the Secure Anonymised Information Linkage (SAIL) Databank. The de-identified data used in this study were made available to accredited researchers only. Those wishing to gain access to the data should contact in the first instance.

Data used in this study are available in the Scottish National Safe Haven (Project Number: 2021-0102), but as restrictions apply they are not publicly available. Access to data may be granted on application to, and subject to approval by, the Public Benefit and Privacy Panel for Health and Social Care (PBPP (https://www.informationgovernance.scot.nhs.uk/pbpphsc/)). Applications are coordinated by eDRIS (electronic Data Research and Innovation Service (https://www.isdscotland.org/Products-and-services/Edris/)). The anonymised data used in this study was made available to accredited researchers only through the Public Health Scotland (PHS) eDRIS User Agreement (https://www.isdscotland.org/Products-and-services/Edris/_docs/eDRIS-User-Agreement-v16.pdf).

The data used in this study are available in the SAIL Databank at Swansea University, Swansea, UK, but as restrictions apply they are not publicly available. All proposals to use SAIL data are subject to review by an independent Information Governance Review Panel (IGRP). Before any data can be accessed, approval must be given by the IGRP. The IGRP gives careful consideration to each project to ensure proper and appropriate use of SAIL data. When access has been granted, it is gained through a privacy protecting safe haven and remote access system referred to as the SAIL Gateway. SAIL has established an application process to be followed by anyone who would like to access data via SAIL at https://www.saildatabank.com/application-process

