## Supplementary figures and tables for "Indirect effects of the first two years of the COVID-19 pandemic on secondary care for cardiovascular disease in the UK: an electronic health record analysis across three countries"

**Supplementary materials**

| **Content** | **Page number** |
| --- | --- |
| Table S1 Coding for cardiovascular disease subtypes and procedures included in the study | 2 |
| Table S2 Demographic characteristics for individuals admitted for cardiovascular disease diagnoses | 3 |
| Table S3 Demographic characteristics for individuals admitted for cardiovascular disease procedures by territory | 5 |
| Figure S1: Study data sources and datasets | 9 |
| Figure S2: Monthly emergency hospital procedures for cardiovascular disease across subtypes, across three countries in the UK and across pre-pandemic (2016-2019) and pandemic (2020-2021) periods. | 10 |
| Figure S3: Monthly elective procedures for cardiovascular disease across subtypes, across three countries in the UK and across pre-pandemic (2016-2019) and pandemic (2020-2021) periods. | 11 |

**Table S1 Codes for cardiovascular disease subtypes and procedures included in the study**

| **Cardiovascular subtypes** | **ICD-10 codes** |
| --- | --- |
| Acute coronary syndrome | I20.0, I21, I22, I24 |
| Heart failure | I50 |
| Acute stroke/ Transient Ischaemic Attack | I60, I61, I63, I64, G45 |
| Aortic Aneurysm | I71 |
| Peripheral arterial disease | I70, I73, I74, I77 |
| Venous thromboembolism | I26, I80, I81, I82 |
| **Cardiovascular Procedures** | **OPSC-4 codes, ICD-10 codes** |
| Percutaneous coronary intervention | K49; K50, K63.1-K63.6; K65; K75 |
| Coronary artery bypass grafts | K40; K41; K42; K43; K44; K45; K46 |
| Pacemaker or cardiac resynchronisation therapy | K59, K60; K61 |
| Ventricular assist device or heart transplant | K01, K02; K54.1; K54.2 |
| Stroke thrombolysis or thrombectomy | I63 AND X83.3, I63 AND L35.4 |
| Carotid endarterectomy or stenting | L29.4; L29.5; L31.4 |
| Cerebral aneurysm coiling | O01 or O02 or O03 or O04 AND Y53 AND Z35 |
| Aortic aneurysm repair | L18; L19; L20; L21; L22; L23; L25; L26; L27; L28 |
| Peripheral limb angioplasty | L54; L63; L66; L71.1 |
| Limb revascularisation, bypass or amputation | L16; L20.6; L21.6; L48; L49; L50; L51; L52; L53; L56; L57; L58; L59; L60; L65.2; L65.3; X09; X10; X11, X12 |
| Pulmonary artery embolectomy or embolisation | L12.4; L12.5; L13 |

ICD-10: international classification of diseases, 10^th^ revision; OPCS-4: Office of population censuses and surveys classification of interventions and procedures, version 4

**Table S2** **– Demographic characteristics for individuals admitted for cardiovascular disease diagnoses**

|  | England | | | Scotland | | Wales | |
| --- | --- | --- | --- | --- | --- | --- | --- |
| Characteristics n (%) | 2016-2019 | 2020-2021 | 2016-2019 | | 2020-2021 | 2016-2019 | 2020-2021 |
| **Cardiac** |  |  |  | |  |  |  |
| ***Acute coronary syndrome*** | |  |  | |  |  |  |
| Men | 276824 (65.2) | 133482 (66.0) | 6576 (63.0) | | 6529 (64.4) | 4339 (63.3) | 4228 (65.0) |
| Age (years) |  |  |  | |  |  |  |
| <50 | 36555 (8.6) | 17541 (8.7) | 825 (7.9) | | 736 (7.3) | 488 (7.1) | 469 (7.2) |
| 50-59 | 75724 (17.8) | 38030 (18.8) | 1974 (18.9) | | 1959 (19.3) | 1174 (17.1) | 1164 (17.9) |
| 60-69 | 94958 (22.4) | 47268 (23.4) | 2492 (23.9) | | 2509 (24.8) | 1588 (23.1) | 1555 (23.9) |
| 70-79 | 105203 (24.8) | 50809 (25.1) | 2522 (24.1) | | 2571 (25.4) | 1797 (26.2) | 1799 (27.7) |
| ≥ 80 | 112427 (26.5) | 48573 (24.0) | 2634 (25.2) | | 2358 (23.3) | 1810 (26.4) | 1514 (23.3) |
| Ethnicity |  |  |  | |  |  |  |
| White | 335062 (78.9) | 155336 (76.8) | 8452 (80.9) | | 8112 (80.1) | 2175 (31.8) | 2338 (36.0) |
| Mixed race | 1728 (0.4) | 933 (0.5) | 14 (0.1) | | 20 (0.1) | <5 | 8 (0.1) |
| Asian | 30599 (7.2) | 14360 (7.1) | 142 (1.4) | | 137 (1.4) | 16 (0.2) | 19 (0.3) |
| Black | 5819 (1.4) | 2795 (1.4) | 11 (0.1) | | 10.5 (0.1) | <5 | <5 |
| Other or unknown | 51659 (12.2) | 28797 (14.2) | 1830 (17.5) | | 1855 (18.3) | 4655 (68.0) | 4125 (63.6) |
| Charlson index |  |  |  | |  |  |  |
| 0 | 158321 (37.3) | 70883 (35.1) | 6176 (59.1) | | 6002 (59.24) | 532 (7.8) | 482 (7.4) |
| 1 | 128510 (30.2) | 59312 (29.3) | 2462 (23.6) | | 2540 (25.1) | 2811 (41.0) | 2520 (38.8) |
| 2 | 65846 (15.5) | 32731 (16.2) | 1062 (10.2) | | 942 (9.3) | 1815 (26.5) | 1818 (28.0) |
| 3+ | 72190 (17.0) | 39295 (19.4) | 748 (7.2) | | 649 (6.4) | 1699 (24.8) | 1682 (25.9) |
| ***Heart failure*** | |  |  | |  |  |  |
| Men | 200885 (54.4) | 102542 (54.4) | 3527 (52.9) | | 3402 (52.5) | 2648 (53.77) | 2473 (53.86) |
| Age (years) |  |  |  | |  |  |  |
| <50 | 11093 (3.0) | 5648 (3.0) | 199 (3.0) | | 214 (3.3) | 130 (2.64) | 122 (2.65) |
| 50-59 | 20883 (5.7) | 11500 (6.1) | 405 (6.1) | | 444 (6.9) | 248 (5.03) | 248 (5.41) |
| 60-69 | 45635 (12.4) | 22697 (12.0) | 951 (14.3) | | 953 (14.7) | 595 (12.09) | 544 (11.85) |
| 70-79 | 96306 (26.1) | 49275 (26.1) | 1861 (27.9) | | 1762 (27.2) | 1364 (27.7) | 1282 (27.9) |
| ≥ 80 | 195281 (52.9) | 99508 (52.8) | 3254 (48.8) | | 3102 (47.9) | 2588 (52.6) | 2395 (52.2) |
| Ethnicity |  |  |  | |  |  |  |
| White | 309677 (83.9) | 156050 (82.7) | 5870 (88.0) | | 5678 (87.7) | 1800 (36.6) | 2072 (45.2) |
| Mixed race | 1418 (0.4) | 769 (0.4) | 11 (0.2) | | 10 (0.2) | <5 | <5 |
| Asian | 21250 (5.8) | 9877 (5.2) | 79 (1.2) | | 91 (1.4) | 12 (0.25) | 20 (0.44) |
| Black | 10035 (2.7) | 5036 (2.7) | 8 (0.1) | | 13 (0.2) | <5 | <5 |
| Other or unknown | 26818 (7.3) | 16896 (9.0) | 703 (10.5) | | 683 (10.6) | 3103 (63.1) | 2487 (54.3) |
| Charlson index |  |  |  | |  |  |  |
| 0 | 74096 (20.1) | 34706 (18.4) | 2508 (37.6) | | 2441 (37.7) | <5 | <5 |
| 1 | 82554 (22.4) | 39697 (21.0) | 1671 (25.1) | | 1725 (26.6) | 1127 (22.9) | 1056 (23.0) |
| 2 | 76921 (20.8) | 38727 (20.5) | 1307 (19.6) | | 1263 (19.5) | 1264 (25.7) | 1206 (26.3) |
| 3+ | 135627 (36.7) | 75498 (40.0) | 1184 (17.8) | | 1046 (16.2) | 2533 (51.4) | 2330 (50.8) |
| **Cerebrovascular** |  |  |  | |  |  |  |
| ***Acute stroke or transient ischaemic attack*** | |  |  | |  |  |  |
| Men | 226362 (50.5) | 116466 (51.7) | 6194 (49.7) | | 6283 (51.1) | 3740 (50.1) | 3644 (51.2) |
| Age (years) |  |  |  | |  |  |  |
| <50 | 31172 (6.9) | 16012 (7.1) | 761 (6.1) | | 689 (5.6) | 449 (6.0) | 426 (6.0) |
| 50-59 | 47446 (10.6) | 25746 (11.4) | 1480 (11.9) | | 1455 (11.8) | 789 (10.6) | 715 (10.0) |
| 60-69 | 73660 (16.4) | 38182 (16.9) | 2390 (19.2) | | 2384 (19.4) | 1278 (17.1) | 1209 (17.0) |
| 70-79 | 118130 (26.3) | 60486 (26.8) | 3410 (27.3) | | 3528 (28.7) | 2067 (27.7) | 2118 (29.8) |
| ≥ 80 | 178156 (39.7) | 84947 (37.7) | 4431 (35.5) | | 4253 (34.6) | 2885 (38.6) | 2652 (37.3) |
| Ethnicity |  |  |  | |  |  |  |
| White | 366735 (81.8) | 177503 (78.8) | 10112 (81.1) | | 10171 (82.6) | 2104 (28.2) | 2415 (34.0) |
| Mixed race | 1601 (0.4) | 1014 (0.4) | 11 (0.08) | | 14 (0.1) | <5 | <5 |
| Asian | 16382 (3.7) | 8847 (3.9) | 101 (0.8) | | 98 (0.8) | 12 (0.2) | 14 (0.2) |
| Black | 9152 (2.0) | 4760 (2.1) | 13 (0.1) | | 17 (0.1) | <5 | 6 (0.1) |
| Other or unknown | 54694 (12.2) | 33249 (14.8) | 2235 (17.9) | | 2008 (16.3) | 5342 (71.6) | 4674 (65.8) |
| Charlson index |  |  |  | |  |  |  |
| 0 | 169443 (37.8) | 75659 (33.6) | 7077 (56.7) | | 6840 (55.6) | 2902 (38.9) | 2793 (39.2) |
| 1 | 110395 (24.6) | 49825 (22.1) | 2805 (22.5) | | 2724 (22.1) | 1831 (24.5) | 1808 (25.4) |
| 2 | 80267 (17.9) | 43724 (19.4) | 1527 (12.3) | | 1643 (13.4) | 1364 (18.3) | 1257 (17.7) |
| 3+ | 88459 (19.7) | 56165 (24.9) | 1063 (8.5) | | 1101 (9.0) | 1370 (18.4) | 1260 (17.7) |
| **Other Vascular** |  |  |  | |  |  |  |
| ***Aortic Aneurysm*** | |  |  | |  |  |  |
| Men | 36433 (76.1) | 14238 (75.1) | 692.(75.9) | | 547.5 (72.5) | 515 (76.5) | 377 (72.9) |
| Age (years) |  |  |  | |  |  |  |
| <50 | 2445 (5.1) | 1090 (5.7) | 33.3 (3.7) | | 34 (4.5) | 25 (3.7) | 30 (5.7) |
| 50-59 | 2918 (6.1) | 1496 (7.9) | 50.3 (5.5) | | 43 (5.7) | 40 (5.9) | 38 (7.5) |
| 60-69 | 9604 (20.1) | 3669 (19.3) | 209 (22.9) | | 168 (22.2) | 134 (19.9) | 103 (19.9) |
| 70-79 | 18567 (38.8) | 7228 (38.1) | 352 (38.6) | | 302 (39.9) | 269 (39.9) | 198 (38.3) |
| ≥ 80 | 14365 (30.0) | 5482 (28.9) | 268 (29.4) | | 209 (27.7) | 206 (30.5) | 148 (28.6) |
| Ethnicity |  |  |  | |  |  |  |
| White | 37482 (78.3) | 14168 (74.7) | 735 (80.6) | | 604 (79.9) | 262 (39.2) | 213 (41.5) |
| Mixed race | 149 (0.3) | 82 (0.4) | <5 | | <5 | <5 | <5 |
| Asian | 629 (1.3) | 298 (1.6) | <5 | | <5 | <5 | <5 |
| Black | 593 (1.2) | 253 (1.3) | <5 | | <5 | <5 | <5 |
| Other or unknown | 9046 (18.9) | 4164 (22.0) | 174 (19.1) | | 147 (19.4) | 407 (60.8) | 300 (58.5) |
| Charlson index |  |  |  | |  |  |  |
| 0 | 19712 (41.2) | 7284 (38.4) | 630 (69.0) | | 481 (63.7) | 286 (42.5) | 206 (39.9) |
| 1 | 14227 (29.7) | 5468 (28.8) | 149 (16.3) | | 151 (20.0) | 186 (27.7) | 165 (31.9) |
| 2 | 7711 (16.1) | 3216 (17.0) | 84 (9.2) | | 74 (9.7) | 98 (14.6) | 78 (15.0) |
| 3+ | 6249 (13.0) | 2997 (15.8) | 50 (5.4) | | 50 (6.6) | 103 (15.3) | 68 (13.3) |
| ***Peripheral arterial disease*** | |  |  | |  |  |  |
| Men | 101842 (58.6) | 42216 (58.7) | 2185 (59.0) | | 1829 (58.9) | 1354 (60.3) | 1169 (63.0) |
| Age (years) |  |  |  | |  |  |  |
| <50 | 19836 (11.4) | 8350 (11.6) | 295 (8.0) | | 248 (8.0) | 188 (8.4) | 128 (6.9) |
| 50-59 | 26569 (15.3) | 10866 (15.1) | 603 (16.3) | | 472 (15.2) | 331 (14.7) | 264 (14.3) |
| 60-69 | 43279 (24.9) | 17609 (24.5) | 961 (25.9) | | 829 (26.7) | 574 (25.6) | 490 (26.4) |
| 70-79 | 48592 (27.9) | 20219 (28.1) | 1090 (29.4) | | 906 (29.2) | 656 (29.2) | 538 (29.0) |
| ≥ 80 | 35628 (20.5) | 14814 (20.6) | 756 (20.4) | | 651 (21.0) | 497 (22.1) | 435 (23.4) |
| Ethnicity |  |  |  | |  |  |  |
| White | 142160 (81.7) | 56427 (78.5) | 3216 (86.8) | | 2632 (84.8) | 806 (36.0) | 793 (43.0) |
| Mixed race | 718 (0.4) | 359 (0.5) | <5 | | <5 | <5 | <5 |
| Asian | 5660 (3.3) | 2515 (3.5) | 24 (0.7) | | 22 (0.7) | <5 | <5 |
| Black | 2912 (1.7) | 1171 (1.6) | <5 | | <5 | <5 | <5 |
| Other or unknown | 22454 (12.9) | 11386 (15.8) | 456 (12.3) | | 445 (14.3) | 1434 (64.0) | 1052 (57.0) |
| Charlson index |  |  |  | |  |  |  |
| 0 | 67397 (38.8) | 24370 (33.9) | 2355 (63.6) | | 1785 (57.5) | 862 (38.4) | 584 (31.5) |
| 1 | 47357 (27.2) | 19098 (26.6) | 626 (16.9) | | 644 (20.7) | 624 (27.8) | 542 (29.2) |
| 2 | 29125 (16.7) | 12946 (18.0) | 445 (12.0) | | 414 (13.3) | 358 (16.0) | 352 (18.9) |
| 3+ | 30025 (17.3) | 15444 (21.5) | 280 (7.6) | | 262 (8.4) | 402 (17.9) | 378 (20.4) |
| ***Venous thromboembolism*** | |  |  | |  |  |  |
| Men | 121772 (48.8) | 69032 (49.9) | 2521 (49.5) | | 2653 (49.3) | 1609 (48.4) | 1864 (49.2) |
| Age (years) |  |  |  | |  |  |  |
| <50 | 57825 (23.2) | 31500 (22.8) | 1220 (24.0) | | 1148 (21.3) | 662 (19.9) | 768 (20.3) |
| 50-59 | 38076 (15.3) | 22682 (16.4) | 774 (15.2) | | 898 (16.7) | 496 (14.9) | 600 (15.8) |
| 60-69 | 47365 (19.0) | 26159 (18.9) | 10255 (20.1) | | 1077 (20.0) | 684 (20.6) | 742 (19.6) |
| 70-79 | 55994 (22.4) | 31391 (22.7) | 1146 (22.5) | | 1297 (24.1) | 808 (24.3) | 940 (24.8) |
| ≥ 80 | 50253 (20.1) | 26536 (19.2) | 926 (18.2) | | 966 (17.9) | 676 (20.3) | 739 (19.5) |
| Ethnicity |  |  |  | |  |  |  |
| White | 210082 (84.2) | 112375 (81.3) | 4177 (82.1) | | 4487 (83.3) | 973 (29.3) | 1416 (37.5) |
| Mixed race | 1393 (0.6) | 884 (0.6) | 6 (0.1) | | 9 (0.2) | <5 | <5 |
| Asian | 6154 (2.5) | 3481 (2.5) | 26 (0.5) | | 19 (0.4) | <5 | 6 (0.1) |
| Black | 6067 (2.4) | 3662 (2.6) | 7 (0.1) | | 12 (0.2) | <5 | <5 |
| Other or unknown | 25817 (10.3) | 17866 (12.9) | 876 (17.2) | | 857 (15.9) | 2344 (70.7) | 2356 (62.4) |
| Charlson index |  |  |  | |  |  |  |
| 0 | 118865 (47.6) | 64012 (46.3) | 2969 (58.3) | | 3051 (56.7) | 1564 (47.0) | 1740 (45.9) |
| 1 | 65512 (26.3) | 34998 (25.3) | 870 (17.1) | | 1013 (18.8) | 744 (22.4) | 856 (22.6) |
| 2 | 35776 (14.3) | 20513 (14.8) | 573 (11.3) | | 609 (11.3) | 423 (12.7) | 480 (12.7) |
| 3+ | 29360 (11.8) | 18745 (13.6) | 680 (13.4) | | 712 (13.2) | 596 (17.9) | 712 (18.8) |

Venous thromboembolism: deep vein thrombosis or pulmonary embolism

**Table S3** **– Demographic characteristics for individuals admitted for cardiovascular disease procedures**

|  | England | | Scotland | | Wales | |
| --- | --- | --- | --- | --- | --- | --- |
| Characteristics n (%) | 2016-2019 | 2020-2021 | 2016-2019 | 2020-2021 | 2016/19 | 2020/21 |
| **Cardiac** |  |  |  |  |  |  |
| ***Percutaneous coronary intervention*** | |  |  |  |  |  |
| Men | 526548 (66.8) | 202595 (68.3) | 12408 (66.5) | 10886 (68.0) | 7319 (65.6) | 6152 (67.4) |
| Age (years) |  |  |  |  |  |  |
| <50 | 76091 (9.7) | 28441 (9.6) | 1810 (9.7) | 1413 (8.8) | 991 (8.9) | 791 (8.7) |
| 50-59 | 161097 (20.5) | 61847 (20.9) | 4266 (22.9) | 3691 (23.1) | 2191 (19.7) | 1772 (19.4) |
| 60-69 | 222606 (28.3) | 83024 (28.0) | 5673 (30.4) | 4835 (30.2) | 3219 (28.9) | 2546 (27.9) |
| 70-79 | 226845 (28.8) | 86264 (29.1) | 4951 (26.6) | 4396 (27.5) | 3344 (30.0) | 2800 (30.7) |
| ≥ 80 | 101079 (12.8) | 36996 (12.5) | 1950 (10.5) | 1676 (10.5) | 1406 (12.6) | 1219 (13.4) |
| Ethnicity |  |  |  |  |  |  |
| White | 597885 (75.9) | 218505 (73.7) | 12990 (69.7) | 10202 (63.7) | 3455 (31.0) | 3035 (33.3) |
| Mixed race | 3849 (0.5) | 1555 (0.5) | 13 (0.1) | 15 (0.1) | 6 (0.1) | 6 (0.1) |
| Asian | 55527 (7.0) | 20461 (6.9) | 223 (1.2) | 175 (1.1) | 30 (0.3) | 32 (0.4) |
| Black | 11134 (1.4) | 4304 (1.5) | 19 (0.1) | 17 (0.1) | 9 (0.1) | <5 |
| Other or unknown | 119323 (15.1) | 51747 (17.4) | 5405 (29.0) | 5602 (35.0) | 7651 (68.6) | 6045 (66.3) |
| Charlson index |  |  |  |  |  |  |
| 0 | 380128 (48.3) | 130839 (44.1) | 12622 (67.7) | 10751 (67.2) | 11054 (99.2) | 9060 (99.3) |
| 1 | 246982 (31.4) | 93287 (31.5) | 4223 (22.6) | 3715 (23.2) | 96 (0.9) | 68 (0.8) |
| 2 | 94263 (12.0) | 40305 (13.6) | 1207 (6.5) | 1042 (6.5) | <5 | <5 |
| 3+ | 66345 (8.4) | 32141 (10.8) | 597 (3.2) | 503 (3.1) | <5 | <5 |
| ***Coronary artery bypass grafts*** | |  |  |  |  |  |
| Men | 50706 (81.5) | 17493 (82.7) | 1054 (80.5) | 837 (80.71) | 810 (79.4) | 537 (82.9) |
| Age (years) |  |  |  |  |  |  |
| <50 | 2865 (4.6) | 1074 (5.1) | 52 (4.0) | 35 (3.4) | 33 (3.2) | 21 (3.2) |
| 50-59 | 10736 (17.3) | 3982 (18.8) | 241 (18.4) | 177 (17.0) | 143 (14.0) | 108 (16.8) |
| 60-69 | 20045 (32.2) | 7056 (33.4) | 467 (35.7) | 388 (37.4) | 320 (31.4) | 220 (34.1) |
| 70-79 | 22765 (36.6) | 7653 (36.2) | 450 (34.3) | 387 (37.3) | 400 (39.2) | 254 (39.2) |
| ≥ 80 | 5793 (9.3) | 1377 (6.5) | 100 (7.6) | 52 (5.0) | 124 (12.1) | 44 (6.7) |
| Ethnicity |  |  |  |  |  |  |
| White | 44977 (72.3) | 14089 (66.6) | 833 (63.6) | 528 (50.9) | 415 (41.0) | 268 (41.6) |
| Mixed race | 238 (0.4) | 100 (0.5) | <5 | <5 | <5 | <5 |
| Asian | 4519 (7.3) | 1437 (6.8) | 15 (1.1) | 10 (0.9) | <5 | <5 |
| Black | 463 (0.7) | 157 (0.7) | <5 | <5 | <5 | <5 |
| Other or unknown | 12007 (19.3) | 5359 (25.3) | 460 (35.2) | 499 (48.1) | 597 (59.0) | 375 (58.4) |
| Charlson index |  |  |  |  |  |  |
| 0 | 22626 (36.4) | 6301 (29.8) | 781 (59.7) | 596 (57.5) | 711 (69.9) | 471 (72.9) |
| 1 | 21093 (33.9) | 6878 (32.5) | 372 (28.4) | 313 (30.2) | 306 (30.1) | 175 (27.1) |
| 2 | 10438 (16.8) | 4335 (20.5) | 106 (8.1) | 91 (8.8) | <5 | <5 |
| 3+ | 8047 (12.9) | 3628 (17.2) | 50 (3.8) | 37 (3.6) | <5 | <5 |
| ***Pacemaker or cardiac resynchronisation therapy*** | | |  |  |  |  |
| Men | 207933 (67.0) | 88009 (67.6) | 2615 (63.8) | 2605 (64.4) | 3036 (65.3) | 2706 (66.7) |
| Age (years) |  |  |  |  |  |  |
| <50 | 22408 (7.2) | 8548 (6.6) | 274 (6.7) | 239.5 (5.9) | 334 (7.2) | 294 (7.2) |
| 50-59 | 29577 (9.5) | 12411 (9.5) | 354 (8.6) | 341 (8.4) | 409 (8.8) | 366 (9.0) |
| 60-69 | 59158 (19.1) | 23955 (18.4) | 702 (17.1) | 713.5 (17.6) | 882 (19.0) | 747 (18.4) |
| 70-79 | 102244 (33.0) | 43064 (33.1) | 1320 (32.2) | 1274 (31.5) | 1585 (34.1) | 1346 (33.2) |
| ≥ 80 | 96791 (31.2) | 42160 (32.4) | 1446 (35.3) | 1478 (36.5) | 1441 (31.0) | 1307 (32.2) |
| Ethnicity |  |  |  |  |  |  |
| White | 251103 (81.0) | 102071 (78.4) | 3231 (78.9) | 3252 (80.4) | 1712 (37.0) | 1434 (35.4) |
| Mixed race | 1151 (0.4) | 543 (0.4) | <5 | <5 | <5 | <5 |
| Asian | 12396 (4.0) | 5042 (3.9) | 28 (0.7) | 28 (0.7) | 11 (0.2) | 8 (0.2) |
| Black | 3874 (1.2) | 1695 (1.3) | <5 | <5 | <5 | <5 |
| Other or unknown | 41654 (13.4) | 20787 (16.0) | 831 (20.29) | 761 (18.8) | 2920 (62.9) | 2609 (64.4) |
| Charlson index |  |  |  |  |  |  |
| 0 | 128245 (41.3) | 50041 (38.5) | 2749 (67.1) | 2715 (67.1) | 3717 (80.0) | 3358 (82.8) |
| 1 | 90461 (29.2) | 36263 (27.9) | 760 (18.6) | 751 (18.6) | 920 (19.8) | 694 (17.1) |
| 2 | 48521 (15.6) | 21771 (16.7) | 365 (8.9) | 382 (9.4) | 10 (0.2) | 6 (0.2) |
| 3+ | 42951 (13.8) | 22063 (17.0) | 222 (5.4) | 199 (4.9) | <5 | <5 |
| ***Ventricular assist device or heart transplant*** | | |  |  |  |  |
| Men | 750 (70.4) | 202 (66.9) | 12 (75.8) | 14 (56.3) | 7 (100) | 6 (100) |
| Age (years) |  |  |  |  |  |  |
| <50 | 528 (49.6) | 162 (53.6) | 9 (56.5) | 11 (45.8) | 6 (100) | <5 |
| 50-59 | 313 (29.4) | 92 (30.5) | 6 (35.5) | 9 (37.5) | 0 | <5 |
| 60-69 | 199 (18.7) | 45 (14.9) | <5 | <5 | 0 | <5 |
| 70-79 | NR | NR | <5 | <5 | 0 | <5 |
| ≥ 80 | NR | NR | <5 | <5 | 0 | <5 |
| Ethnicity |  |  |  |  |  |  |
| White | 778 (73.1) | 228 (75.5) | 6 (38.7) | 13 (52.1) | 8 (100) | 6 (100) |
| Mixed race | NR | NR | NR |  | 0 | 0 |
| Asian | 95 (8.9) | 23 (7.6) | <5 | <5 | 0 | 0 |
| Black | 38 (3.6) | 6 (2.0) | NR |  | 0 | 0 |
| Other or unknown | 134 (12.6) | 41 (13.6) | 9 (59.7) | 12 (47.9) | 0 | 0 |
| Charlson index |  |  |  |  |  |  |
| 0 | 362 (34.0) | 124 (41.1) | 9 (59.7) | 17 (70.8) | 11 (100) | 8 (100) |
| 1 | 306 (28.7) | 80 (26.5) | <5 | 6 (22.9) | 0 | 0 |
| 2 | 179 (16.8) | 46 (15.2) | <5 | <5 | 0 | 0 |
| 3+ | 218 (20.5) | 52 (17.2) | <5 | <5 | 0 | 0 |
| **Cerebrovascular** |  |  |  |  |  |  |
| ***Stroke thrombolysis or thrombectomy*** | | |  |  |  |  |
| Men | 8592 (55.6) | 4978 (57.3) | <5 | <5 | 139 (57.4) | 152 (58.2) |
| Age (years) |  |  |  |  |  |  |
| <50 | 1121 (7.3) | 711 (8.2) | <5 | <5 | 15 (6.2) | 12 (4.7) |
| 50-59 | 1912 (12.4) | 1220 (14.1) | <5 | <5 | 28 (11.3) | 30 (11.7) |
| 60-69 | 2914 (18.9) | 1714 (19.7) | <5 | <5 | 48 (19.6) | 52 (20.0) |
| 70-79 | 4515 (29.2) | 2578 (29.7) | <5 | <5 | 73 (30.2) | 80 (31.3) |
| ≥ 80 | 4990 (32.3) | 2460 (28.3) | <5 | <5 | 79 (32.7) | 84 (32.4) |
| Ethnicity |  |  |  |  |  |  |
| White | 12281 (79.5) | 6384 (73.5) | <5 | <5 | 84 (34.8) | 92 (35.2) |
| Mixed race | 52 (0.3) | 40 (0.5) | <5 | <5 | <5 | <5 |
| Asian | 508 (3.3) | 320 (3.7) | <5 | <5 | <5 | <5 |
| Black | 216 (1.4) | 122 (1.4) | <5 | <5 | <5 | <5 |
| Other or unknown | 2395 (15.5) | 1817 (20.9) | <5 | <5 | 158 (65.2) | 170 (64.8) |
| Charlson index |  |  |  |  |  |  |
| 0 | 5984 (38.7) | 2672 (30.8) | <5 | <5 | 245 (100) | 265 (100) |
| 1 | 3374 (21.8) | 1540 (17.7) | <5 | <5 | 0 | 0 |
| 2 | 3181 (20.6) | 2259 (26.0) | <5 | <5 | 0 | 0 |
| 3+ | 2913 (18.9) | 2212 (25.5) | <5 | <5 | 0 | 0 |
| ***Carotid endarterectomy or stenting*** | | |  |  |  |  |
| Men | 9366 (67.5) | 3839 (68.8) | 274 (68.7) | 209 (68.2) | 149 (65.8) | 78 (64.3) |
| Age (years) |  |  |  |  |  |  |
| <50 | 281 (2.0) | 144 (2.6) | 7 (1.8) | 7 (2.3) | <5 | <5 |
| 50-59 | 1520 (11.0) | 653 (11.7) | 58 (14.6) | 48 (15.5) | 30 (13.4) | 12 (9.6) |
| 60-69 | 3575 (25.8) | 1550 (27.8) | 128 (32.0) | 88 (28.6) | 68 (30.4) | 34 (28.8) |
| 70-79 | 5530 (39.9) | 2149 (38.5) | 143 (35.8) | 117 (38.2) | 88 (39.8) | 53 (44.2) |
| ≥ 80 | 2965 (21.4) | 1081 (19.4) | 63 (15.9) | 48 (15.5) | 36 (16.4) | 21 (17.5) |
| Ethnicity |  |  |  |  |  |  |
| White | 10469 (75.5) | 3958 (71.0) | 300 (75.3) | 224 (72.9) | 87 (38.5) | 52 (43.0) |
| Mixed race | 26 (0.2) | 8 (0.1) | <5 | <5 |  |  |
| Asian | 342 (2.5) | 133 (2.4) | <5 | <5 |  |  |
| Black | 43 (0.3) | 23 (0.4) | <5 | <5 |  |  |
| Other or unknown | 2991 (21.6) | 1455 (26.1) | 96 (24.1) | 82 (26.6) | 139 (61.5) | 69 (57.0) |
| Charlson index |  |  |  |  |  |  |
| 0 | 5961 (43.0) | 2279 (40.9) | 270 (67.8) | 202 (65.9) | 225 (100) | 122 (100) |
| 1 | 4073 (29.4) | 1534 (27.5) | 76 (19.06) | 64 (20.7) | 0 | 0 |
| 2 | 2168 (15.6) | 952 (17.1) | 33 (8.2) | 31 (10.0) | 0 | 0 |
| 3+ | 1669 (12.0) | 812 (14.6) | 20 (5.1) | 11 (3.4) | 0 | 0 |
| ***Cerebral aneurysm coiling*** | | |  |  |  |  |
| Men | 1921 (30.4) | 1007 (31.1) | 68 (27.3) | 79 (29.6) | 44 (30.5) | 38 (29.5) |
| Age (years) |  |  |  |  |  |  |
| <50 | 1741 (27.6) | 774 (23.9) | 71 (28.7) | 63 (23.5) | 40 (28.3) | 26 (20.8) |
| 50-59 | 1916 (30.4) | 1003 (31.0) | 84 (34.0) | 81 (30.5) | 51 (36.3) | 41 (32.8) |
| 60-69 | 1597 (25.3) | 845 (26.1) | 58 (23.4) | 76 (28.6) | 34 (23.8) | 41 (32.8) |
| 70-79 | 880 (13.9) | 512 (15.8) | 29 (11.5) | 38 (14.1) | 16 (11.6) | 17 (13.6) |
| ≥ 80 | 179 (2.8) | 100 (3.1) | 6 (2.4) | 9 (3.2) | <5 | <5 |
| Ethnicity |  |  |  |  |  |  |
| White | 4553 (72.1) | 2204 (68.2) | 149 (60.3) | 167 (62.7) | 47 (33.2) | 28 (22.3) |
| Mixed race | 37 (0.6) | 28 (0.9) | <5 | <5 | <5 | <5 |
| Asian | 190 (3.0) | 124 (3.8) | <5 | <5 | <5 | <5 |
| Black | 174 (2.8) | 87 (2.7) | <5 | <5 | <5 | <5 |
| Other or unknown | 1359 (21.5) | 791 (24.5) | 94 (38.1) | 97 (36.53 | 94 (66.8) | 98 (77.7) |
| Charlson index |  |  |  |  |  |  |
| 0 | 4125 (65.3) | 1971 (60.9) | 218 (88.0) | 234 (88.0) | 142 (100) | 126 (100) |
| 1 | 1352 (21.4) | 745 (23.0) | 22 (8.7) | 22 (8.1) | 0 | 0 |
| 2 | 586 (9.3) | 340 (10.5) | 7 (2.9) | 9 (3.2) | 0 | 0 |
| 3+ | 250 (4.0) | 178 (5.5) | <5 | <5 | 0 | 0 |
| **Other vascular** |  |  |  |  |  |  |
| ***Aortic aneurysm repair*** | |  |  |  |  |  |
| Men | 36084 (73.3) | 12305 (74.9) | 1046 (71.7) | 786 (72.2) | 558 (72.8) | 388 (74.9) |
| Age (years) |  |  |  |  |  |  |
| <50 | 5244 (10.6) | 1975 (12.0) | 126 (8.6) | 105 (9.7) | 73 (9.5) | 58 (11.2) |
| 50-59 | 4194 (8.5) | 1654 (10.1) | 144 (9.8) | 113 (10.4) | 62 (8.1) | 50 (9.7) |
| 60-69 | 10564 (21.4) | 3480 (21.2) | 374 (25.7) | 282 (25.9) | 164 (21.4) | 108 (20.7) |
| 70-79 | 18233 (37.0) | 6114 (37.2) | 550 (37.7) | 424 (39.0) | 285 (37.2) | 199 (38.3) |
| ≥ 80 | 11017 (22.4) | 3215 (19.6) | 266 (18.2) | 164 (15.1) | 182 (23.8) | 104 (20.0) |
| Ethnicity |  |  |  |  |  |  |
| White | 38115 (77.4) | 11988 (72.9) | 994 (68.1) | 698 (64.1) | 315 (41.6) | 222 (43.2) |
| Mixed race | 206 (0.4) | 78 (0.5) | <5 | <5 | <5 | <5 |
| Asian | 1339 (2.7) | 445 (2.7) | 8 (0.5) | 8 (0.7) | <5 | <5 |
| Black | 553 (1.1) | 181 (1.1) | <5 | <5 | <5 | <5 |
| Other or unknown | 9039 (18.4) | 3746 (22.8) | 457 (31.3) | 379 (34.8) | 443 (58.5) | 293 (56.8) |
| Charlson index |  |  |  |  |  |  |
| 0 | 21802 (44.3) | 6696 (40.7) | 1024 (70.2) | 749 (68.8) | 670 (87.6) | 386 (74.4) |
| 1 | 14712 (29.9) | 4849 (29.5) | 283 (19.4) | 211 (19.4) | 95 (12.4) | 133 (25.7) |
| 2 | 7263 (14.7) | 2636 (16.0) | 104 (7.1) | 93 (8.5) | <5 | <5 |
| 3+ | 5475 (11.1) | 2257 (13.7) | 48 (3.3) | 37 (3.4) | <5 | <5 |
| ***Peripheral limb angioplasty*** | | |  |  |  |  |
| Men | 61640 (67.5) | 24672 (68.4) | 1272 (63.8) | 1097 (63.9) | 663 (64.5) | 489 (67.2) |
| Age (years) |  |  |  |  |  |  |
| <50 | 3792 (4.2) | 1392 (3.9) | 103 (5.2) | 83 (4.8) | 41 (4.0) | 28 (3.9) |
| 50-59 | 12277 (13.4) | 4668 (12.9) | 307 (15.4) | 246 (14.3) | 137 (13.3) | 82 (11.2) |
| 60-69 | 24685 (27.0) | 9642 (26.7) | 547 (27.42) | 468 (27.2) | 275 (26.8) | 196 (27.0) |
| 70-79 | 29822 (32.7) | 11988 (33.2) | 628 (31.5) | 553 (32.2) | 332 (32.3) | 242 (33.2) |
| ≥ 80 | 20720 (22.7) | 8390 (23.3) | 410 (20.5) | 369 (21.5) | 242 (23.6) | 180 (24.7) |
| Ethnicity |  |  |  |  |  |  |
| White | 74565 (81.7) | 28354 (78.6) | 1690 (84.7) | 1379 (80.2) | 414 (40.4) | 317 (43.9) |
| Mixed race | 281 (0.3) | 126 (0.3) | <5 | <5 | <5 | <5 |
| Asian | 1906 (2.1) | 767 (2.1) | 10 (0.5) | 14 (0.8) | <5 | <5 |
| Black | 1303 (1.4) | 588 (1.6) | <5 | <5 | <5 | <5 |
| Other or unknown | 13241 (14.5) | 6245 (17.3) | 289 (14.5) | 323 (18.8) | 610 (59.6) | 404 (56.1) |
| Charlson index |  |  |  |  |  |  |
| 0 | 30942 (33.9) | 9703 (26.9) | 1277 (64.0) | 921 (53.6) | 1014 (98.7) | 702 (97.3) |
| 1 | 25033 (27.4) | 9345 (25.9) | 302 (15.1) | 348 (20.3) | 13 (1.3) | 20 (2.8) |
| 2 | 16904 (18.5) | 7362 (20.4) | 281 (14.1) | 314 (18.3) |  |  |
| 3+ | 18417 (20.2) | 9670 (26.8) | 136 (6.8) | 136 (7.9) |  |  |
| ***Limb revascularisation, bypass or amputation*** | | |  |  |  |  |
| Men | 48386 (70.8) | 20393 (73.5) | 1877 (69.0) | 1731 (70.7) | 1048 (70.8) | 945 (75.2) |
| Age (years) |  |  |  |  |  |  |
| <50 | 8938 (13.1) | 3427 (12.4) | 304 (11.2) | 248 (10.1) | 168 (11.4) | 123 (9.8) |
| 50-59 | 12231 (17.9) | 5141 (18.5) | 521 (19.2) | 453 (18.5) | 251 (16.9) | 214 (17.1) |
| 60-69 | 17374 (25.4) | 7336 (26.5) | 758 (27.9) | 720 (29.4) | 396 (26.7) | 377 (30.0) |
| 70-79 | 18587 (27.2) | 7605 (27.4) | 772 (28.4) | 698 (28.5) | 422 (28.5) | 344 (27.4) |
| ≥ 80 | 11202 (16.4) | 4226 (15.2) | 365 (13.4) | 328 (13.4) | 244 (16.5) | 198 (15.7) |
| Ethnicity |  |  |  |  |  |  |
| White | 56527 (82.7) | 21977 (79.2) | 2331 (85.7) | 2008 (82.1) | 597 (40.5) | 572 (45.8) |
| Mixed race | 229 (0.3) | 139 (0.5) | <5 | <5 | <5 | <5 |
| Asian | 1293 (1.9) | 575 (2.1) | 19 (0.7) | 17 (0.7) | <5 | <5 |
| Black | 912 (1.3) | 383 (1.4) | <5 | <5 | <5 | <5 |
| Other or unknown | 9371 (13.7) | 4661 (16.8) | 366 (13.4) | 417 (17.0) | 879 (59.5) | 678 (54.2) |
| Charlson index |  |  |  |  |  |  |
| 0 | 19657 (28.8) | 6691 (24.1) | 1522 (56.0) | 1198 (49) | 1472 (99.4) | 1248 (100) |
| 1 | 16691 (24.4) | 6030 (21.7) | 422 (15.5) | 465 (19.0) | 10 (0.6) | 0 |
| 2 | 15848 (23.2) | 7097 (25.6) | 551 (20.3) | 567 (23.2) | 0 | 0 |
| 3+ | 16136 (23.6) | 7917 (28.5) | 224 (8.2) | 217 (8.9) | 0 | 0 |
| ***Pulmonary artery embolectomy or embolisation*** | | |  |  |  |  |
| Men | 1938 (52.0) | 774 (52.6) | 58 (46.7) | 39 (50.7) | 27 (49.54) | 18 (61.4) |
| Age (years) |  |  |  |  |  |  |
| <50 | 2512 (67.4) | 958 (65.1) | 57 (45.7) | 36 (46.1) | 36 (85.8) | 15 (100) |
| 50-59 | 329 (8.8) | 125 (8.5) | 16 (13.1) | 15 (19.5) | <5 | <5 |
| 60-69 | 375 (10.1) | 170 (11.5) | 23 (18.7) | 12 (14.9) | 6 (14.2) | <5 |
| 70-79 | 383 (10.3) | 159 (10.8) | 20 (15.7) | 11 (13.6) | <5 | <5 |
| ≥ 80 | 127 (3.4) | 60 (4.1) | 9 (6.8) | <5 | <5 | <5 |
| Ethnicity |  |  |  |  |  |  |
| White | 2577 (69.2) | 1049 (71.3) | 86 (69.4) | 47 (61.0) | 35 (69.2) | 13 (44.1) |
| Mixed race | 69 (1.9) | 25 (1.7) | <5 | <5 | <5 | <5 |
| Asian | 301 (8.1) | 137 (9.3) | <5 | <5 | <5 | <5 |
| Black | 112 (3.0) | 28 (1.9) | <5 | <5 | <5 | <5 |
| Other or unknown | 667 (17.9) | 233 (15.8) | 37 (29.8) | 29 (37.0) | 16 (30.9) | 16 (55.9) |
| Charlson index |  |  |  |  |  |  |
| 0 | 1809 (48.6) | 700 (47.6) | 77 (62.0) | 57 (73.4) | 53 (100) | 30 (100) |
| 1 | 1327 (35.6) | 567 (38.5) | 24 (19.1) | 11 (14.3) | 0 | 0 |
| 2 | 338 (9.1) | 122 (8.3) | 14 (11.5) | <5 | 0 | 0 |
| 3+ | 252 (6.8) | 83 (5.6) | 9 (7.4) | <5 | 0 | 0 |

**Figure S1: Study data sources and datasets**

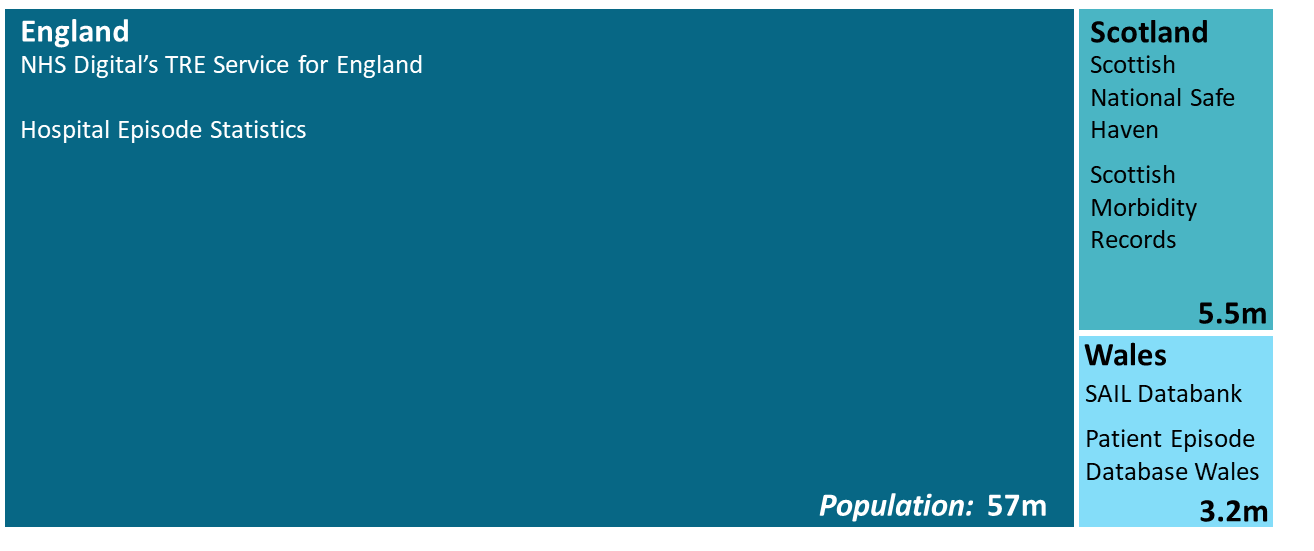

TRE: trusted research environment

**Figure S2: Monthly emergency hospital procedures for cardiovascular disease across subtypes, across three countries in the UK and across pre-pandemic (2016-2019) and pandemic (2020-2021) periods.**

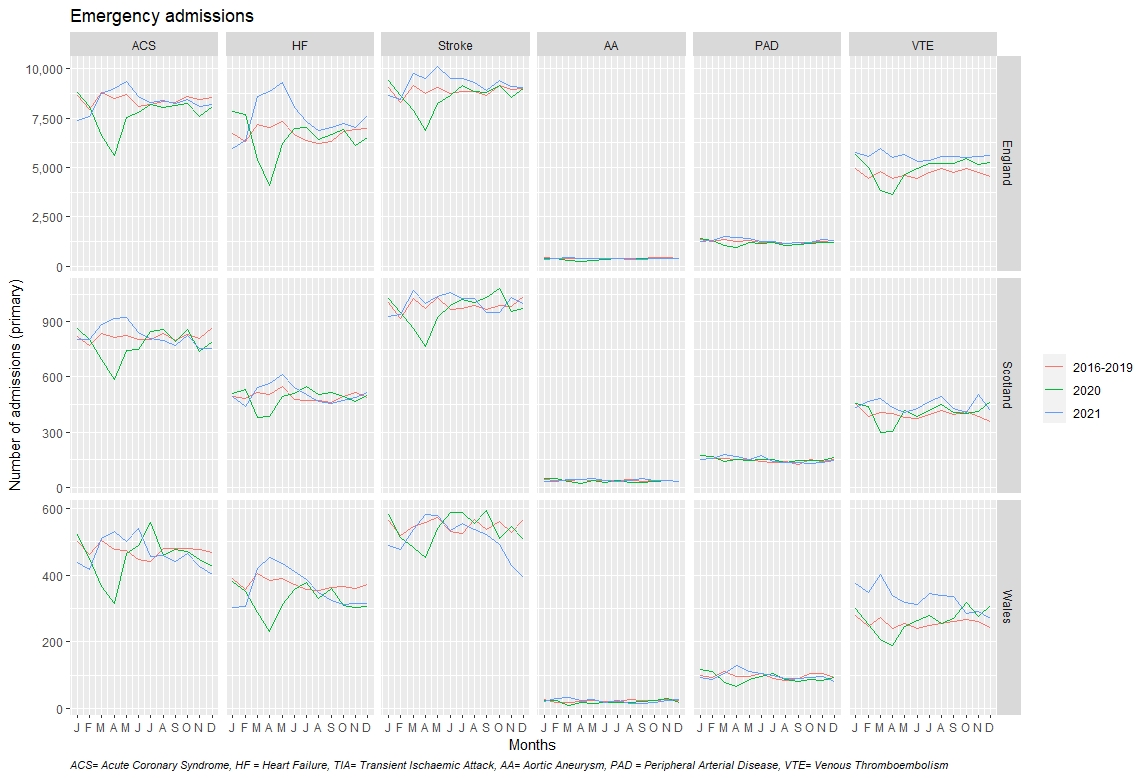

**Figure S3: Monthly elective procedures for cardiovascular disease across subtypes, across three countries in the UK and across pre-pandemic (2016-2019) and pandemic (2020-2021) periods.**

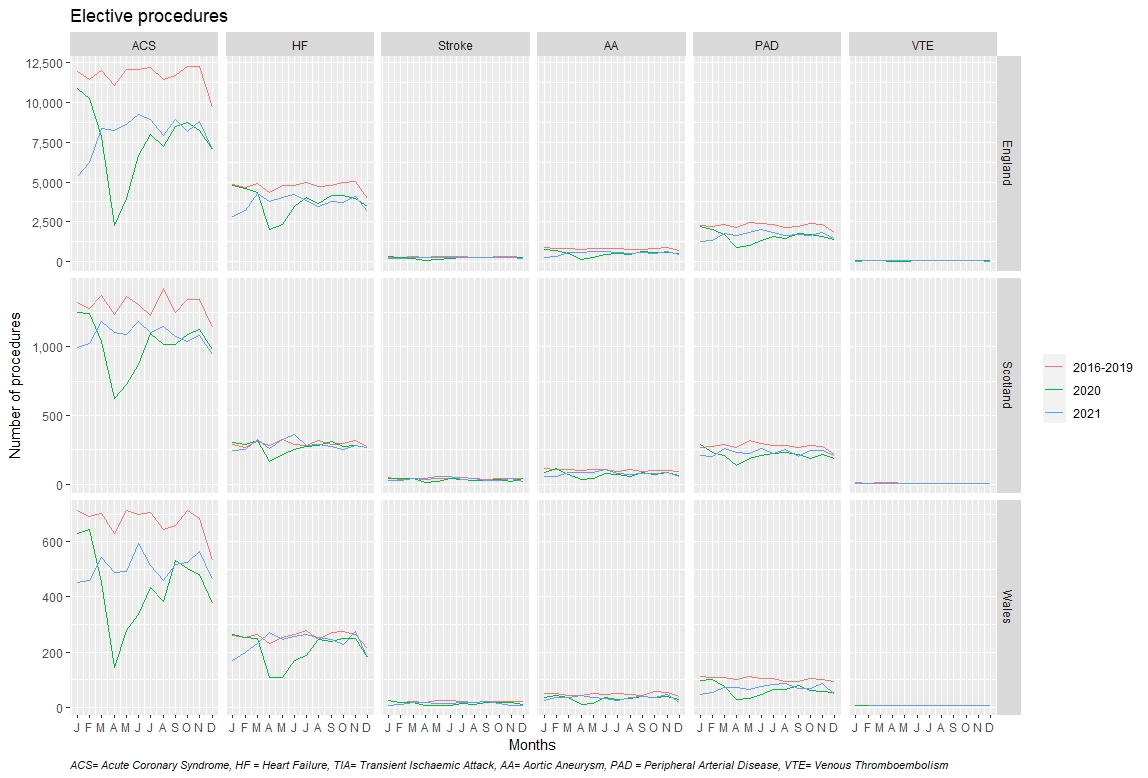
